# Determinants of COPD Stage Progression and Regression: a Markov Transition Analysis of The COPDGene Cohort

**DOI:** 10.1101/2025.01.17.25320745

**Authors:** Luz M. Sánchez-Romero, Andrew F. Brouwer, Rafael Meza, David T. Levy, Rossana Torres-Alvarez, MeiLan K. Han

**Author notes:** Corresponding author: Luz M Sánchez-Romero, Georgetown University Medical Center 2115 Wisconsin Ave, NW, Suite 300, Washington, DC 20007. **Author contributions:** L.M.S-R takes responsibility for the data analysis and content of this manuscript. L.M.S-R, A.F.B, R.M, M.K.H designed the study. M.K.H, A.F.B, R.T-A contributed to the acquisition, statistical analysis, or interpretation of the data. L.M.S-R and A.F.B drafted the manuscript. A.F.B, R.M, D.T.L, M.K H contributed to the critical revision of the manuscript for important intellectual content. **Financial/nonfinancial disclosure:** None declared.

## Abstract

**Rationale:** Chronic obstructive pulmonary disease (COPD) is a leading cause of death but with variable progression.

**Objective:** Estimate factors influencing transition rates between PRISm and GOLD stages.

**Methods:** Using a Markov multistate model, transition rates between GOLD-0, PRISm and GOLD-1, GOLD-2 and GOLD 3-4 were estimated for 5,728 US adult ever cigarette users from the COPDGene cohort over 10-years. We calculated one and five-year transition probabilities for progressive and regressive transitions and estimated the mean sojourn time for severity states.

**Main Results:** GOLD-1 and PRISm individuals spent the least time in any single stage (GOLD-1: 6 years; PRISm: 7 years). PRISm and GOLD-1 individuals were equally likely to transition to GOLD-2 vs. GOLD-0 (PRISm: HR 1.09, 95% confidence interval [CI] 0.90-1.33, GOLD-1 (HR 1.15, 95%CI 0.93-1.42) per five-year period, but rarely transition between PRISm and GOLD-1. Individuals at GOLD-0 were equally likely to progress to GOLD-1 or PRISm (HR 1.11, 95%CI 0.93-1.31) but the transient time for this stage was the longest of any GOLD stage (16 years, 95%CI 15.2-17.3). GOLD-2 was the most likely stage to progress (HR 2.4, 95%CI 1.9-3.02) to GOLD 3-4 vs. regress to GOLD-1. For GOLD-2 individuals, current smoking status (HR 0.84, 95%CI 0.67-1.06) or intensity (HR 0.84, 95%CI 0.54-1.29) was not associated with disease progression.

**Conclusions:** GOLD-1 and PRISm are the most transient stages equally likely to regress to GOLD-0 or progress to GOLD-2 and may benefit from smoking cessation interventions. GOLD-2 individuals are the most likely to progress and may benefit most from targeted disease interventions.

---

Chronic obstructive pulmonary disease (COPD) is a deadly, progressive disease that accounts for roughly three million deaths annually.^1^ COPD term encompasses lung conditions that cause persistent and progressive airflow limitation, including emphysema and chronic bronchitis.^2^ Despite this, COPD remains severely underdiagnosed, and diagnosis is often delayed until more severe airflow obstruction is present.^3–5^

COPD is defined by a forced expiratory volume and forced vital capacity (FVC) ratio< 0.70. COPD disease staging and treatment are based on the severity of a patient’s airflow limitation, measured by spirometry. COPD stages are defined using the forced expiratory volume in 1 second (FEV_1_) and the forced vital capacity (FVC).^6^ The Global Initiative for Chronic Obstructive Lung Disease (GOLD) classification suggests four stages based on the FEV_1_/FVC ratio, ranging from GOLD 1 (mild airflow limitation) to GOLD 4 (very severe limitation).^7^ In recent years, clinical research has also demonstrated the significance of another stage, known as preserved ratio impaired spirometry (PRISm), accounting for a reduced lung function not captured by the traditional COPD definition.^8,9^

Even in its early stages (PRISm or GOLD 1), the disease has been associated with respiratory symptoms, as well as increased coronary heart disease risk^10^, lung cancer^11^ and mortality.^12–14^ Detecting the disease at early stages allows patients and healthcare providers to address risk factors (smoking, in particular), slow progression, and improve prognosis.^14–16^ While some studies have assessed the longitudinal trajectories of COPD stages,^12,17–20^ few have modeled the specific transitions from/to less to/from more severe disease stages. A better understanding of the disease’s early stages characteristics and the determinants of COPD progression and regression is needed.

In this paper, we used the COPDGene study cohort to estimate the underlying transition rates between airflow limitation categories (PRISm and GOLD) for US adults who have ever smoked to identify determinants of disease transition at each stage as well as the time spent in each severity stage. Our analysis aims to provide insights into how patient characteristics influence their propensity to progress or regress across COPD severity stages.

## STUDY DESIGN and METHODS

### Data and variable definitions

We used data from the COPDGene study, a US observational longitudinal cohort of the underlying factors of COPD.^21^ COPDGene includes 10,192 current and former smokers (≥10 pack-years), aged 45-80 years old with and without COPD diagnosis. Data were collected at baseline (phase-1: 2008-2011) with two follow-ups at five-year intervals (phase-2: 2012-2016 and phase-3: 2018-2021). We restricted our analysis to individuals with information on tobacco use and post-bronchodilator spirometry values (FEV_1_ and FVC) in at least two phases.

In addition to the spirometry and cigarette use (current and former) information, we used data on gender (male, female), age group (39-54, 55-64, 65-90 years), race (Non-Hispanic White [NHW], Non-Hispanic Black [NHB]), cigarette pack-years (<20 pack-years, ≥20 pack-years), body mass index (BMI) (normal: ≤24.9 kg/m^2^, overweight: 25-29.9 kg/m^2^, obesity: ≥30 kg/m^2^), and previous diagnosis of COPD (yes, no). Previous diagnosis of COPD variable was defined as those who said *yes* or *don’t know* to “Have you ever had COPD” to overcome recall bias and *yes* to “was it diagnosed by doctor or other health professional?” or “In the past 12 months, have you received medical treatment, taken medications or used and inhaler for COPD?”

We removed participants with missing information for stage or any covariate in any given phase and then removed individuals with only one remaining observation. Our final analytical sample comprised 5,728 individuals with 13,259 observations (phase-1: 5,715, phase-2: 5,584, phase-3: 1,960) (Figure 1).

**Figure 1.**
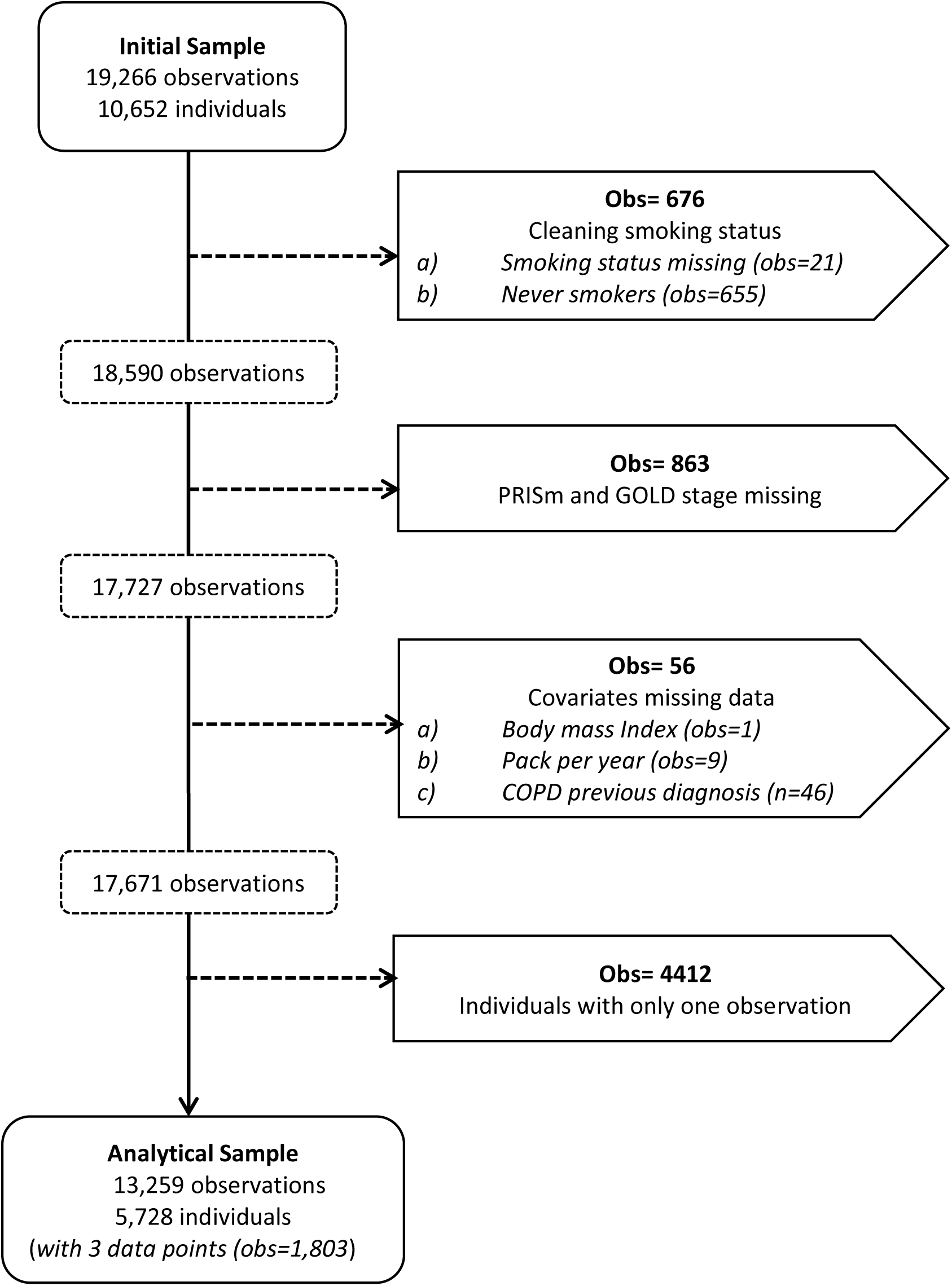
Exclusion Criteria for COPDGene analytical sample: Phases 1 to 3.

We categorized each individual in each phase into one of six disease severity stages based on post-bronchodilator FEV_1_/FVC values, using the Global Initiative for Chronic Obstructive Lung Disease (GOLD) classification (FEV_1_/FVC ratio and predicted FEV_1_)^22^: GOLD 0 (i.e., normal spirometry) (≥0.7 and ≥80%); PRISm (≥0.7 and <80%); GOLD 1 (<0.7 and ≥80%); GOLD 2 (<0.7 and 50%≤ to <80%); and GOLD 3-4 (<0.7 and <50%). We included death as a sixth stage.

### Model reduction

Estimates of rates for rare transitions can be unreliable and uninformative, and their inclusion in a multistate transition model can interfere with estimating other parameters. Thus, we identified state transitions that were rare enough that they should be considered negligible based on the COPDGene data (Figure 2). COPD is a progressive chronic disease; we assumed that individuals could only change their FEV_1_ or FEV_1_/FVC categories one step at a time and not change both metrics simultaneously. Individuals can still progress between phases, e.g., from GOLD 0 to GOLD 2, but the model implicitly assumes that they have an intermediate state that we do not necessarily observe due to the sparse temporal sampling (e.g., GOLD 1 or PRISm in this example).

**Figure 2.**
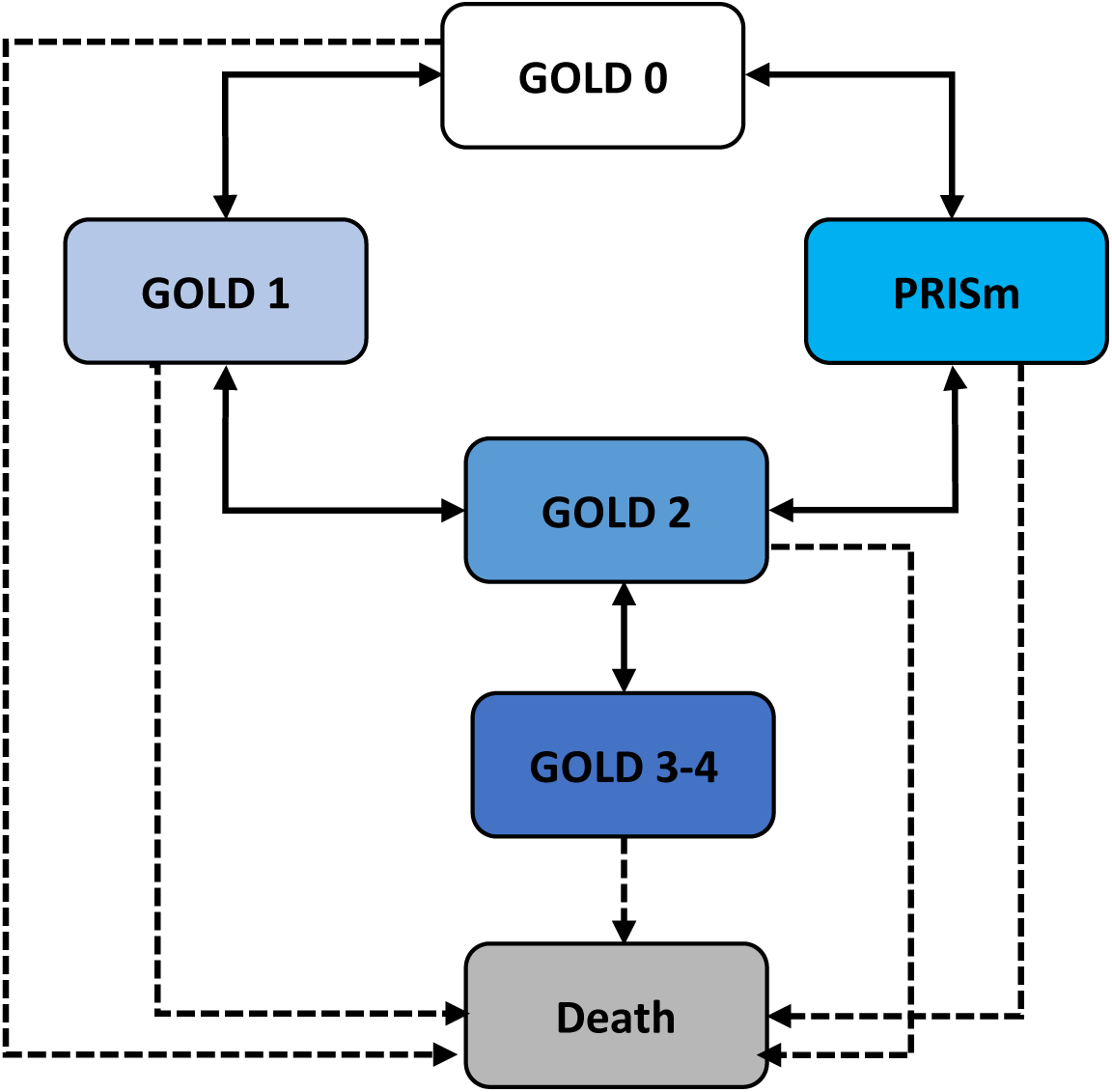
Direct transitions allowed between PRISm and chronic obstructive pulmonary (COPD) GOLD severity stages.

### Transition modeling

We used a univariate Markov multistate transition model to estimate the annual transition hazard rates and covariate hazard ratios (HR) between stages (GOLD 0, PRISm, GOLD 1, GOLD 2, GOLD 3-4, death) overall and for each sociodemographic characteristic (defined at each participant’s most recent phase) in univariable models. A Markov multistate transition model is a continuous-time, finite-state stochastic process that assumes that transition rates depend only on the current state without the influence of transition history.^23^ This model estimates the instantaneous risk of transitions from one health stage to another (transitions hazard rates).

These transition hazard rates and ratios can be used to compute the probabilities of any individual being in each stage after a specified period. We calculated the expected transition probabilities for individuals in each stage after one year and five years.^24^ Additionally, we estimated transition hazard ratios for pairs of transitions to estimate the relative propensity of progression or regression at the early stages of the disease. Specifically, we compared the transition hazard rates for PRISm to GOLD 2 vs to GOLD 0, for GOLD 1 to GOLD 2 vs. to GOLD 0; for GOLD 2 to GOLD 3-4 vs. to GOLD 1 or to PRISm, for GOLD 0 to GOLD1 vs. to PRISm, and for GOLD 1 or PRISm to GOLD 2. Finally, we estimated the mean sojourn time in a single stay in each severity state using the *sojourn.msm* function of the msm-package for R. All analyses were performed using the COPDGene-Phenotype Dataset without sampling variability in the pulmonary function testing. Although the spirometry measurements are only observed every five years, the model accounts for the possibility and likelihood of transitions among unobserved, intermediate states when estimating the transition rates, hazard ratios, and mean time in state.

### Sensitivity Analysis

Assignment of participants to an airflow limitation category depends on whether one uses pre- or post-bronchodilator (BD) values.^25,26^ We assessed whether using pre-BD spirometry values to define stages impacted the estimates of COPD transitions (Supplementary Appendix 1).

## RESULTS

### Descriptive

Our study included 5,738 adults (age 38-90, mean 59.5±8.6 years) at baseline (phase-1). At baseline 50.5% were male, 69.5% NHW, 73.1% BMI≥25kg/m^2^, and 73.0% had no previous COPD diagnosis. At baseline, both current (49.2%) and former (50.8%) smokers were equally represented, with 85.3% exceeding 20 pack-years (average 43.3±23.6). Severity stages prevalence at baseline were: 47.5% GOLD 0, 8.9% GOLD 1, 19.3% GOLD 2, 12.0% GOLD 3-4, and 12.4% PRISm (Table 1). As shown in Figure-E2, PRISm individuals mostly remained stable or transitioned to GOLD 0 or GOLD 2, while GOLD 1 individuals more often progressed to GOLD 2. Transitions between PRISm and GOLD 1 were uncommon.

**Table 1.** Descriptive characteristics of the analytic sample by study phase.

|  | Phase<br>1 | N=571<br>5 | Phase<br>2 | N=558<br>4 | Phase<br>3 | N=196<br>0 | Total | N=132<br>59 |
| --- | --- | --- | --- | --- | --- | --- | --- | --- |
| **Gender** |  |  |  |  |  |  |  |  |
| Male | 2886 | 50.5 | 2813 | 50.38 | 971 | 49.54 | 6670 | 50.31 |
| Female | 2829 | 49.5 | 2771 | 49.62 | 989 | 50.46 | 6589 | 49.69 |
| **Age group**<br>**(years)** |  |  |  |  |  |  |  |  |
| 39 to 54 | 930 | 16.27 | 885 | 15.85 | 357 | 18.21 | 2172 | 16.33 |
| 55 to 64 | 3146 | 55.05 | 3074 | 55.05 | 1111 | 56.68 | 7331 | 55.34 |
| 65 to 90 | 1639 | 28.68 | 1625 | 29.1 | 492 | 25.1 | 3756 | 28.33 |
| **Race** |  |  |  |  |  |  |  |  |
| NH-White | 3947 | 69.06 | 3890 | 69.66 | 1374 | 70.1 | 9211 | 69.47 |
| NH-Black | 1768 | 30.94 | 1694 | 30.34 | 586 | 29.9 | 4048 | 30.53 |
| **Cigarette use** |  |  |  |  |  |  |  |  |
| Current | 2812 | 49.2 | 2181 | 39.06 | 657 | 33.52 | 5650 | 42.61 |
| Former | 2903 | 50.8 | 3403 | 60.94 | 1303 | 66.48 | 7609 | 57.39 |
| **Cigarette**<br>**pack years** |  |  |  |  |  |  |  |  |
| less than 20 | 839 | 14.68 | 715 | 12.8 | 228 | 11.63 | 1782 | 13.44 |
| 20 or more | 4876 | 85.32 | 4869 | 87.2 | 1732 | 88.37 | 11477 | 86.56 |
| **Body Mass**<br>**Index (kg/m2)** |  |  |  |  |  |  |  |  |
| Normal<br>(≤24.9) | 1539 | 26.93 | 1547 | 27.7 | 567 | 28.93 | 3653 | 27.55 |
| Overweight<br>(25 to 29.9) | 1975 | 24.56 | 1915 | 34.29 | 708 | 36.12 | 4598 | 34.68 |
| Obesity (≥30) | 2201 | 38.51 | 2122 | 38 | 685 | 34.95 | 5008 | 37.77 |
| **COPD**<br>**diagnosis** |  |  |  |  |  |  |  |  |
| No | 4171 | 72.98 | 4068 | 72.85 | 1481 | 75.56 | 9720 | 73.31 |
| Yes | 1544 | 27.02 | 1516 | 27.15 | 479 | 24.44 | 3539 | 26.69 |
| **COPD-GOLD**<br>**Severity Stage** |  |  |  |  |  |  |  |  |
| Gold 0 | 2712 | 47.45 | 2415 | 43.25 | 815 | 41.58 | 5942 | 44.81 |
| PRISm | 706 | 12.35 | 696 | 12.46 | 231 | 11.79 | 1633 | 12.32 |
| Gold 1 | 507 | 8.87 | 528 | 9.46 | 238 | 12.14 | 1273 | 9.6 |
| Gold 2 | 1102 | 19.28 | 1118 | 20.02 | 383 | 19.54 | 2603 | 19.63 |
| Gold 3-4 | 688 | 12.04 | 827 | 14.81 | 293 | 14.95 | 1808 | 16.64 |
| Death | - | - | - | - | - | - | 964 | 6.75 |

NH-White: Non-Hispanic White; NH-Black- Non-Hispanic Black

### Transitions between PRISm & GOLD

We estimated the one- and five-year transition probabilities averaged over the three phases of data (Figure 3A, 3B). The underlying transition rates and probabilities are given in Table-E3. The least persistent stage was GOLD 1, with 84.8% and 45.8% of GOLD 1 participants remaining after one and five years, respectively. Similarly, 86.5% and 50.3% of the PRISm participants remained in PRISm after one and five years. GOLD 3-4 participants had the highest probability of dying after one (6.4%) and five years (27.3%). Followed by individuals at GOLD 2 with a 13.8% probability of dying after five years. The mean sojourn time in a single state followed a similar pattern with individuals spending around 6 years (95%CI 5.45, 6.61y) at GOLD 1, 7 years (95%CI 6.25, 7.47y) at PRISm and 8 years (95%CI 7.51, 8.68y) at GOLD 2 before transitioning to other stages (Figure 4).

**Figure 3.**
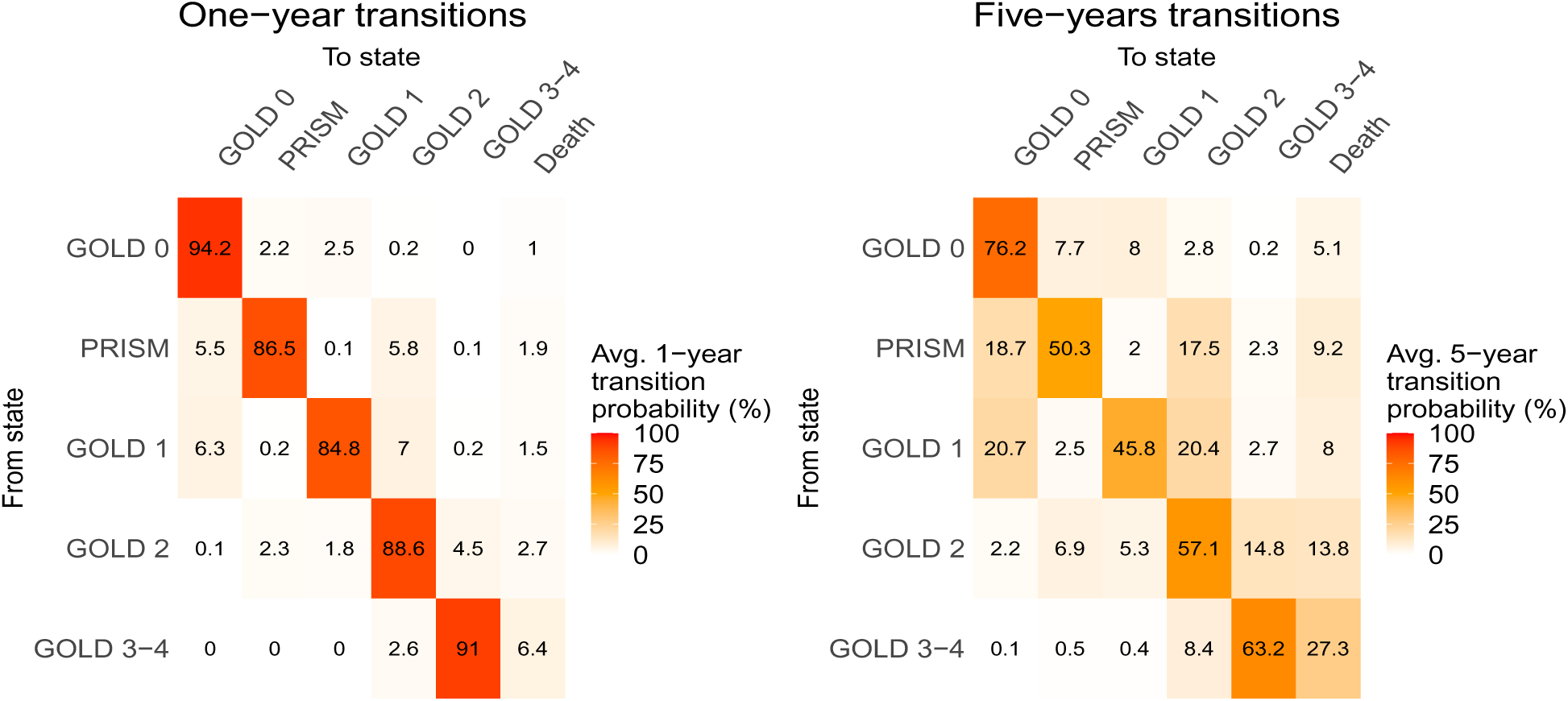
One-year (A) and five-year (B) transition probabilities between PRISm, GOLD severity stages and death, estimated from all-wave sample.

**Figure 4.**
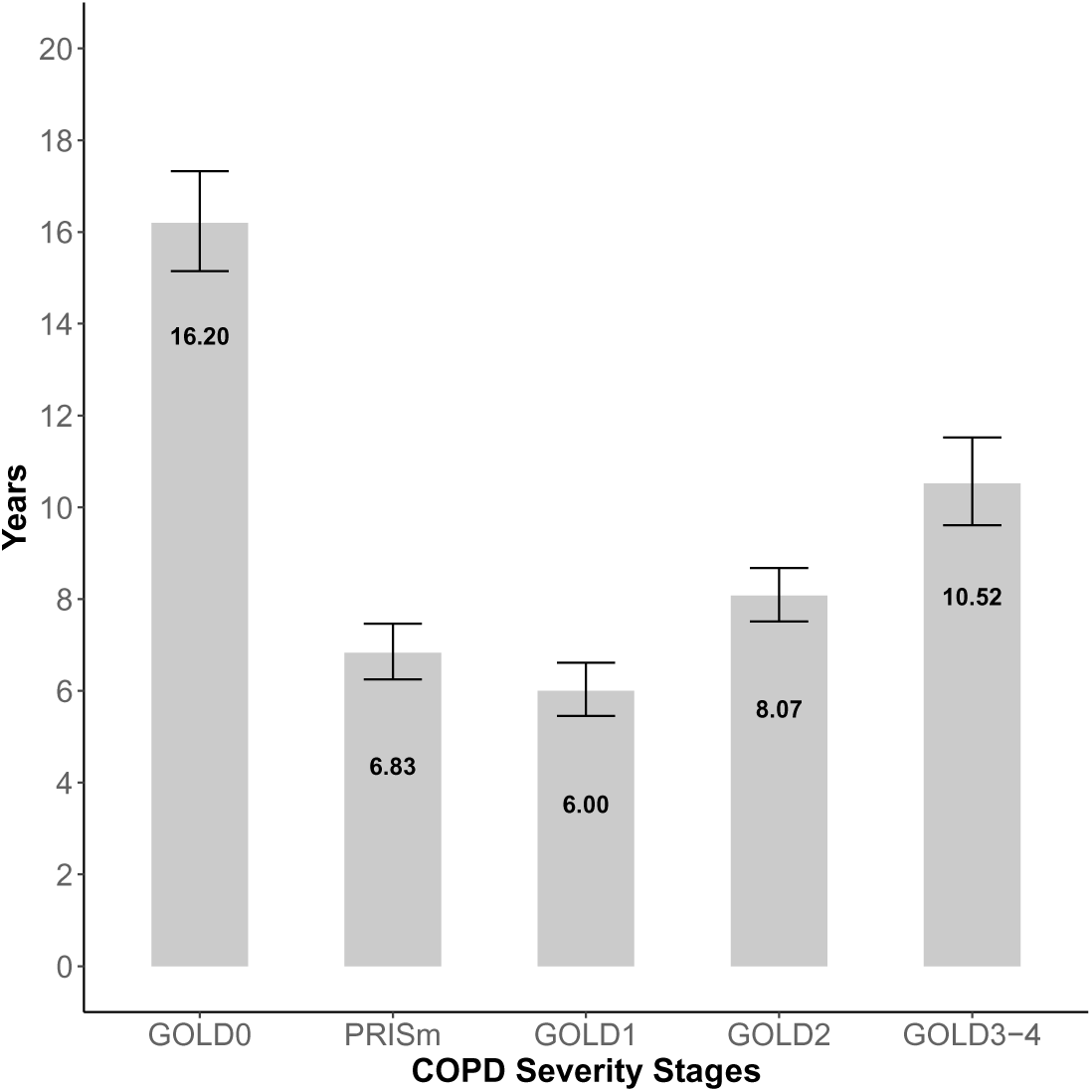
Estimated univariable mean sojourn time of years spend in a single COPD severity stage before transitioning.

We evaluated the relative rates of transitions between GOLD severity stages for individuals at earlier stages of the disease (GOLD 0, PRISm, GOLD 1, and GOLD 2). Individuals at GOLD 0 have similar odds to progressing to GOLD 1 or PRISm. Similarly, no differences in odds to progress to GOLD 2 was observed when coming from GOLD 1 or PRISm. Individuals with a mild severity of obstruction were more likely to progress than to regress to a less severe stage than. Individuals at GOLD 2 were more likely to progress to GOLD 3-4 than to regress to GOLD 1 (HR 2.40 95%CI 1.91, 3.02) or PRISm (HR 1.90, 95%CI 1.54, 2.34) and they were equally likely to regress to GOLD 1 or PRISm (HR 0.79, 95%CI 0.60, 1.05) (Table 2).

**Table 2.** Hazard ratios (univariable) and 95% confidence intervals of the comparison regression to progression hazard rates for the GOLD early disease stages.

| Demographic | From GOLD 0 to<br>GOLD 1 vs.<br>PRISm (ref) | From PRISm to<br>GOLD 2 vs.<br>GOLD 0 (ref) | From GOLD 1 to<br>GOLD 2 vs.<br>GOLD 0(ref) | From GOLD 1 to<br>GOLD 2<br>vs. PRISm to<br>GOLD 2 (ref) | From GOLD 2 to<br>GOLD3-4 vs.<br>PRISm (ref) | From GOLD 2 to<br>GOLD 3-4 vs.<br>GOLD 1 (ref) | From GOLD 2 to<br>GOLD 1 vs.<br>PRISm (ref) |
| --- | --- | --- | --- | --- | --- | --- | --- |
| **Overall** | 1.11<br>(0.93, 1.31) | 1.09<br>(0.90, 1.33) | 1.15<br>(0.93, 1.42) | 1.22<br>(1.00, 1.50) | **1.90**<br>**(1.54, 2.34)<sup>b</sup>** | **2.40**<br>**(1.91, 3.02)<sup>b</sup>** | 0.79<br>(0.60, 1.05) |
| **Gender** |  |  |  |  |  |  |  |
| Male | 1.20<br>(0.94,1.53) | **1.38**<br>**(1.03,1.86)<sup>b</sup>** | 1.34<br>(1.00,1.79) | 1.02<br>(0.76,1.35) | **2.35**<br>**(1.73,3.20)<sup>b</sup>** | **2.50**<br>**(1.82,3.42)<sup>b</sup>** | 0.94<br>(0.63,1.42) |
| Female | 1.03<br>(0.82,1.31) | 0.91<br>(0.70,1.18) | 0.97<br>(0.72,1.31) | **1.49**<br>**(1.11,2.00)<sup>b</sup>** | **1.58**<br>**(1.19,2.10)<sup>b</sup>** | **2.28**<br>**(1.62,3.19)<sup>b</sup>** | 0.69<br>(0.46,1.03) |
| **Age group (years)** |  |  |  |  |  |  |  |
| 39 to 54 | 0.88<br>(0.58,1.35) | 1.10<br>(0.69,1.76) | 1.24<br>(0.63,2.45) | 1.74<br>(0.93,3.25) | 0.95<br>(0.53,1.68) | 2.19<br>(0.97,4.95) | 0.43<br>(0.17,1.07) |
| 55 to 64 | 1.03<br>(0.82,1.28) | 0.89<br>(0.70,1.14) | 1.16<br>(0.87,1.54) | 1.13<br>(0.87,1.46) | **1.65**<br>**(1.27,2.15)<sup>b</sup>** | **3.50**<br>**(2.38,5.15)<sup>b</sup>** | **0.47**<br>**(0.30,0.74)<sup>a</sup>** |
| 65 to 90 | **1.51**<br>**(1.01,2.27)<sup>b</sup>** | **1.89**<br>**(1.20,2.97)** | 1.16<br>(0.82,1.65) | **0.45**<br>**(0.32,0.64)<sup>a</sup>** | **2.71**<br>**(1.81,4.05)<sup>b</sup>** | **1.77**<br>**(1.27,2.47)<sup>b</sup>** | 1.53<br>(0.98,2.39) |
| **Race** |  |  |  |  |  |  |  |
| Non-Hispanic<br>White | **1.56**<br>**(1.25,1.94)<sup>b</sup>** | 1.21<br>(0.94,1.57) | 1.20<br>(0.95,1.52) | 1.14<br>(0.89,1.46) | **2.02**<br>**(1.57,2.60)<sup>b</sup>** | **2.13**<br>**(1.65,2.77)<sup>b</sup>** | 0.95<br>(0.68,1.31) |
| Non-Hispanic<br>Black | **0.67**<br>**(0.50,0.89)<sup>a</sup>** | 0.92<br>(0.68,1.25) | 1.01<br>(0.64,1.60) | 1.45<br>(0.98,2.14) | **1.59**<br>**(1.10,2.29)<sup>b</sup>** | **3.42**<br>**(2.04,5.73)<sup>b</sup>** | **0.46**<br>**(0.25,0.85)<sup>a</sup>** |
| **Cigarette use** |  |  |  |  |  |  |  |
| Current | 0.97<br>(0.77,1.22) | 1.20<br>(0.92,1.57) | **1.52**<br>**(1.11,2.08)<sup>b</sup>** | **1.57**<br>**(1.19,2.05)<sup>b</sup>** | **1.73**<br>**(1.29,2.32)<sup>b</sup>** | **3.11**<br>**(2.10,4.58)<sup>b</sup>** | **0.56**<br>**(0.35,0.89)<sup>a</sup>** |
| Former | **1.31**<br>**(1.02,1.67)<sup>b</sup>** | 0.94<br>(0.71,1.26) | 0.88<br>(0.66,1.18) | 0.94<br>(0.69,1.29) | **2.02**<br>**(1.51,2.70)<sup>b</sup>** | **2.04**<br>**(1.52,2.73)<sup>b</sup>** | 0.99<br>(0.69,1.42) |
| **Cigarette pack**<br>**years** |  |  |  |  |  |  |  |
| less than 20 | 0.83<br>(0.55,1.27) | 0.67<br>(0.39,1.13) | **0.30**<br>**(0.13,0.67)<sup>a</sup>** | 0.71<br>(0.30,1.68) | 0.94<br>(0.52,1.71) | **2.77**<br>**(1.16,6.61)<sup>b</sup>** | **0.34**<br>**(0.13,0.88)<sup>a</sup>** |
| 20 or more | 1.18<br>(0.98,1.42) | 1.19<br>(0.96,1.47) | **1.31**<br>**(1.05,1.64)<sup>b</sup>** | 1.23<br>(1.00,1.52) | **2.08**<br>**(1.66,2.60)<sup>b</sup>** | **2.38**<br>**(1.87,3.02)<sup>b</sup>** | 0.88<br>(0.65,1.18) |
| **Body Mass Index (kg/m<sup>2</sup>)** |  |  |  |  |  |  |  |
| Normal (≤ 24.9 ) | **1.78**<br>**(1.28,2.49)<sup>b</sup>** | 1.40<br>(0.87,2.26) | 1.42<br>(1.00,2.02) | 1.00<br>(0.68,1.48) | **3.70**<br>**(2.35,5.84)<sup>b</sup>** | **3.06**<br>**(2.01,4.65)<sup>b</sup>** | 1.21<br>(0.67,2.19) |
| Overweight (25 to 29.9) | **1.89**<br>**(1.40,2.56)<sup>b</sup>** | 1.41<br>(0.96,2.08) | 1.02<br>(0.73,1.43) | 0.97<br>(0.69,1.37) | **1.61**<br>**(1.11,2.35)<sup>b</sup>** | **1.72**<br>**(1.18,2.52)<sup>b</sup>** | 0.94<br>(0.57,1.54) |
| Obesity (≥30) | **0.51**<br>**(0.38,0.69)<sup>a</sup>** | 0.92<br>(0.71,1.20) | 0.95<br>(0.62,1.44) | **1.53**<br>**(1.04,2.26)<sup>b</sup>** | **1.40**<br>**(1.02,1.93)<sup>b</sup>** | **2.47**<br>**(1.67,3.66)<sup>b</sup>** | **0.57**<br>**(0.37,0.88)<sup>a</sup>** |
| **COPD diagnosis** |  |  |  |  |  |  |  |
| No | 1.05<br>(0.88,1.26) | 0.88<br>(0.70,1.10) | 1.01<br>(0.80,1.27) | **1.37**<br>**(1.08,1.75)<sup>b</sup>** | 1.02<br>(0.77,1.36) | 0.30<br>(0.14,0.63) | 0.83<br>(0.59,1.17) |
| Yes | **1.78**<br>**(1.06,2.99)<sup>b</sup>** | **2.17**<br>**(1.38,3.41)<sup>b</sup>** | **2.35**<br>**(1.30,4.25)<sup>b</sup>** | 0.96<br>(0.65,1.43) | **3.31**<br>**(2.36,4.64)<sup>b</sup>** | * | 0.65<br>(0.38,1.11) |

a: Statistical significant hazard ratio <1 (Regression to a less severe stage is more likely/reference group)
b: Statistical significant hazard ratio ≥1 (Progression to a more severe stage is more likely)
\*Value insufficient power.

### Transition hazard ratios by sociodemographic group

All covariate hazard ratios and transition probabilities matrices are given in Figures E3 (1-7) and Tables E4-5.

### Progression

Figure 5 shows the covariate HRs related to disease progression. A previous COPD diagnosis was consistently associated with progression at all stages. The influence of other sociodemographic variables was dependent on the initial GOLD stage. There were no differences by gender in the rates of progression for any category. Individuals aged 65-90 had greater progression rates from GOLD 0 to GOLD 1 (HR 1.75; 95%CI 1.38, 2.24) and PRISm to GOLD 2 (HR 1.39 95%CI 1.00, 1.92) than 50–64-year-olds. NHB participants were more likely to progress from GOLD 0 to PRISm (HR 1.79; 95%CI 1.42, 2.26) and GOLD 2 to GOLD 3-4 (HR 1.29 95 %CI 1.01, 1.66) than NHW participants.

**Figure 5.**
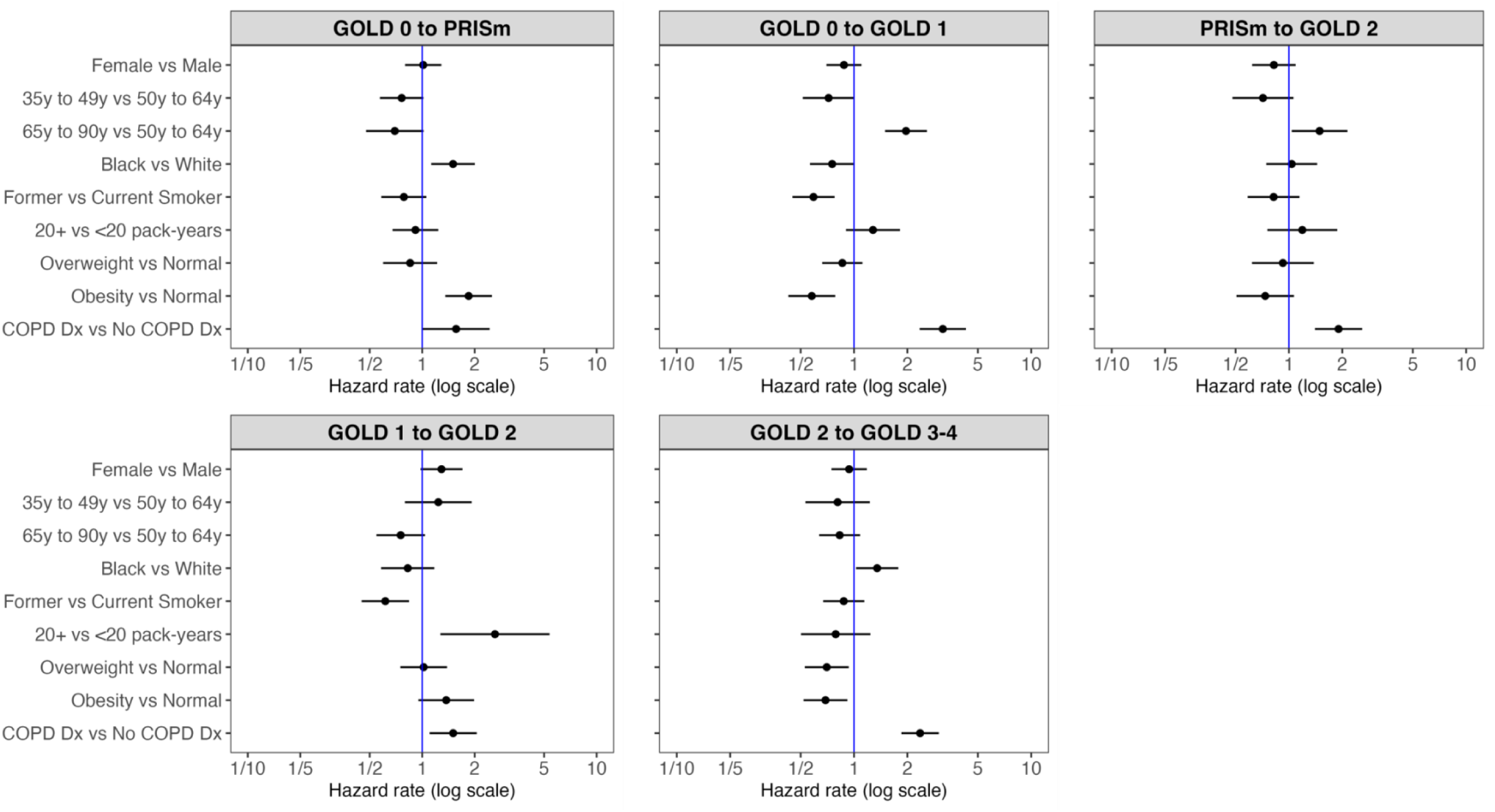
HRs by sociodemographic characteristics for transitions corresponding to COPD progression to a more severe GOLD stage.

Smoking status and intensity influenced transitions at early disease stages. Individuals that reported cigarette use of 20+ pack-years were more likely to progress from GOLD 1 to GOLD 2 (HR 2.52 95%CI 1.24, 5.11) than those with <20 pack-years. Those who previously smoked were less likely than individuals who currently smoked to progress from GOLD 0 to PRISm (HR 0.64; 95%CI 0.51, 0.81) and from GOLD 1 to GOLD 2 (HR 0.61; 95%CI 0.46, 0.80).

There were no consistent patterns between body mass index (BMI) and transitions: individuals with obesity were more likely to progress from GOLD 0 to PRISm (HR 1.85, 95%CI 1.37, 2.49) but less likely to progress from GOLD 0 to GOLD 1 (HR 0.53, 95%CI 0.39, 0.71) than individuals with normal BMI. This was also observed with individuals with a BMI≥25kg/m^2^ being less likely to progress from GOLD 2 to GOLD 3-4 (overweight [HR 0.67 95%CI 0.51,0.88] and obesity [HR 0.69 95%CI 0.53, 0.91]) than individuals with normal BMI.

### Regression

Figure 6 shows the HRs related to disease regression. No sociodemographic characteristic was consistently associated with disease regression in all categories, but there were recurring patterns. We observed that for multiple transitions (GOLD 1 to GOLD 0, GOLD 2 to PRISm, and GOLD 3-4 to GOLD 2), women had greater regression rates than men (HR 1.67 [95%CI 1.22, 2.29] for GOLD 1 to GOLD 0; HR 1.46 [95 %CI 1.02, 2.09]) for GOLD 2 to PRISm; and HR 1.58 [95%CI 1.09, 2.27) for GOLD 3-4 to GOLD 2) and that individuals with obesity had greater regression rates than individuals with normal BMI (HR 1.72 (95%CI 1.14, 2.57) for GOLD 1 to GOLD 0, HR 1.82 (95%CI 1.12, 2.97) for GOLD 2 to PRISm; and HR 1.75 (95%CI 1.13, 2.72) for GOLD 3-4 to GOLD 2).

**Figure 6.**
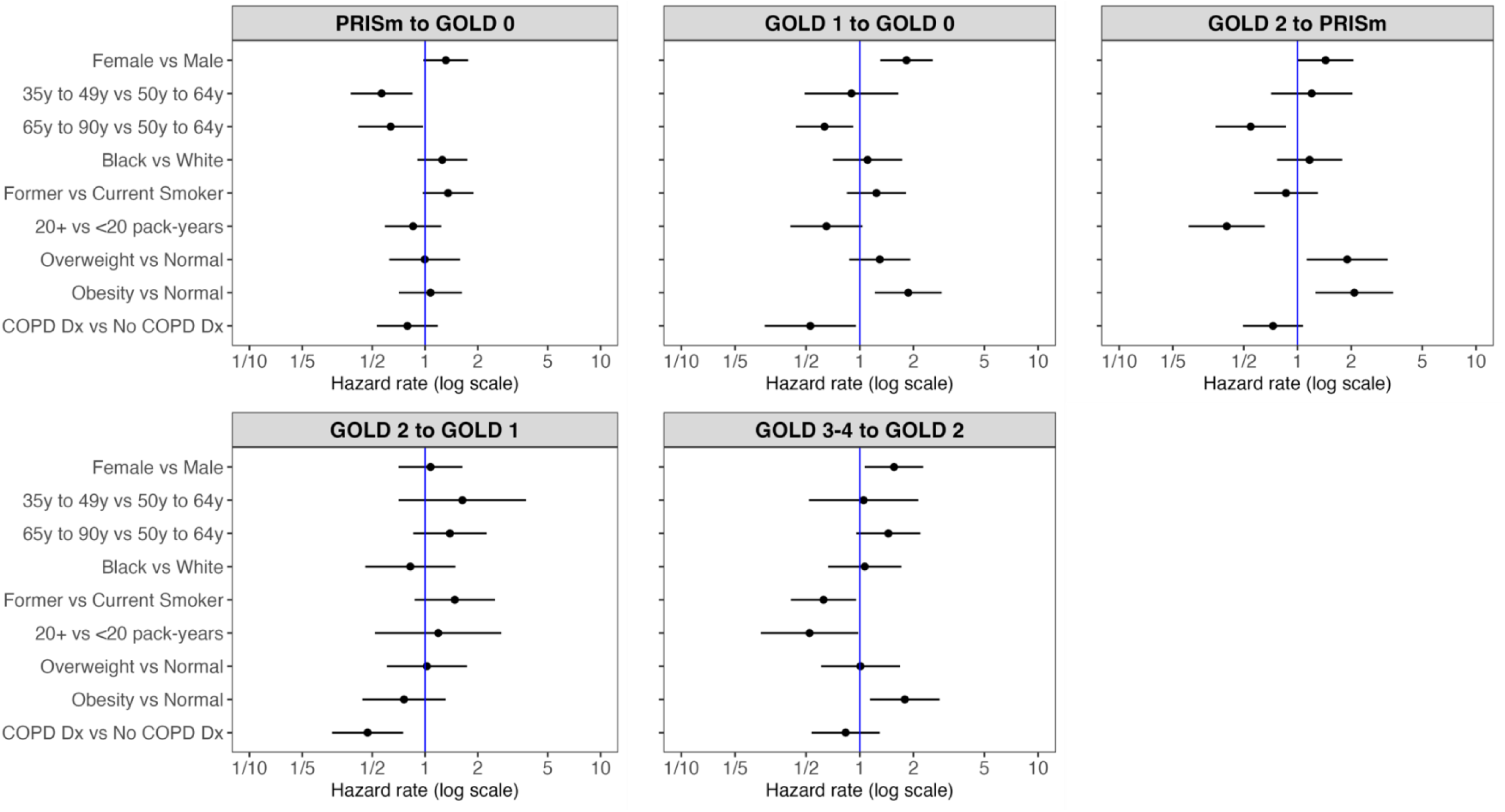
HRs by sociodemographic characteristics for transitions corresponding to COPD progression to a less severe GOLD stage.

Patients 65-90 years old were generally less likely to regress than patients ages 50-64 years (HR 0.66 (95%CI 0.44, 0.97) for PRISm to GOLD 0, HR 0.70 (95%CI 0.50, 0.99) for GOLD 1 to GOLD 0, and HR 0.49 (95%CI 0.32, 0.74) for GOLD 2 to PRISm). NHB participants were more likely than NHW participants to regress from GOLD 2 to PRISm (HR 1.65, 95%CI 1.42, 240).

Individuals smoking 20+ pack-years were less likely to regress vs. those smoking <20 pack-years from GOLD 1 to GOLD 0 (HR 0.57; 95%CI 0.37, 0.88), from GOLD 2 to PRISm (HR 0.38; 95%CI 0.23, 0.61) and GOLD 3-4 to GOLD 2 (HR 0.53; 95%CI 0.29, 0.97). However, being a former smoker was associated with a lower rate of regression from GOLD 3-4 to GOLD 2 than being a current smoker (HR 0.66, 95%CI 0.46, 0.96).

Additionally, individuals with previous COPD diagnosis were less likely than those without a diagnosis to regress from GOLD 2 to PRISm (HR 0.65; 95%CI 0.45, 0.95) or from GOLD 2 to GOLD 1 (HR 0.51; 95%CI 0.32, 0.81).

## DISCUSSION

In a cohort of US ever-smoking adults, we found that GOLD 1 and PRISm were the most dynamic disease stages, with <50% of participants remaining in GOLD 1 and PRISm after five years and a mean sojourn time of around 6.4 years in those stages. Intriguingly, we also found similar likelihoods of these individuals progressing to more severe disease stages versus regressing to less severe ones during that period. The associations of sociodemographic variables with COPD transitions generally depended on the disease stage. A previous COPD diagnosis was associated with disease progression at all stages but had the biggest association with transitions from GOLD 2 to GOLD 3-4

Our study results are consistent with a prior report by Wan et al.,^27^ also using the COPGene Cohort. Wan reported that, around 50% (51.9%-53.9%) of the sample transitioned in and out of PRISm within five years compared to ≥86% that remained in an obstructive state. Here, we reported that after five years, only with 50.3% of the sample stayed at PRISm and only 45.8% remained at GOLD 1. Similar results have also been observed in other populations. Soriano et al.,^19^ using a cohort of Spanish population, assessed the transitions between GOLD 2017 severity stages A, B, C and D (airflow limitations and assessment of symptoms and risk of exacerbations) within five years. The study reported that Grade A (COPD+ 01-exacerbation history and mMRC0-1 CAT <10) was the most frequent and stable stage and that regression from B to A was more likely within the first year. These results were not directly comparable to our study, as we only considered FEV_1_/FVC values, but it supports our conclusion that early stages of the disease are the most dynamic stages.

COPD is considered a progressive disease without a cure. Nevertheless, our study shows that COPD patients can regress to a less severe stage, especially individuals at GOLD 1 and PRISm. It is possible that early lung injury may resolve, particularly among the PRISm group. Regression may also represent a certain degree of bronchodilator reversibility or test variability. Across the various models, risk factors associated with regression included female gender, lower smoking pack-years, obesity, and no prior physician diagnosis of COPD. Other studies have reported this regression phenomenon.^19,27–29^ We conducted a sensitivity analysis using pre-BD spirometry values for GOLD classification to investigate this more in-depth. Our analysis did not find any significant difference in transition rates compared to applying post-BD values for classification. However during our study, we only considered spirometry-defined COPD, without other variables that assess the inflammatory process or that function as disease progression markers such as treatment, exacerbations, biomarkers, or CT imaging.^30^ Spirometry values may vary day-to-day and thus the appearance of regression in our study could be a result of an overly pessimistic initial test or an excessively optimistic later test. This concern is mitigated somewhat by including three phases of data and complemented by a complete-case transition analysis with 1803 individuals with three phases measures, which show no significant differences in transition estimates compared to the main analysis (Figures E5-E4). Still, more evidence needs to be collected to characterize and investigate the determinants of regression to less severe GOLD stages.

COPD progression is also highly influenced by other individual and environmental factors.^31,32^ From our analysis, we observed that the pattern of influence on transitioning within COPD varies between GOLD severity stages, supporting the thesis of COPD’s heterogeneous presentation.^33^ Like Wan et al., we observed that NHB participants had a higher rate of transitions than NHW participants, particularly for progressing or regression from PRISm at early stages. Detection, categorization (i.e., restrictive or non-restrictive) and treatment of PRISm is important due to the increased morbidity and mortality risk.^12,34–36^ However, the literature has reported that NHB individuals have a higher probability of underdiagnosis,^37^ than the NHW population.

Our results showed that BMI (≥30 kg/m^2^) was associated with progression from GOLD 0 to PRISm but with a lower progression rate to more severe stages. It is important to study high BMI’s impact on PRISm transition, which has also been reported by other studies^28,38^ as PRISm diagnosis has been associated with cardiovascular disease risks. Furthermore, low BMI (<20kg/m^2^) has also been reported as a risk factor for COPD incidence and progression, severity of exacerbations and higher risk mortality.^39–41^ Due to a limited sample size (less than 5%) of observations with BMI <20 kg/m^2^), analysis of this subgroup was not feasible for this study. Further research with a larger sample is warranted to explore this population.

Current smoking and pack-years were also associated with the progression of the disease at early stages from GOLD 1 to GOLD 2 but not PRISm. A history of smoking was associated with a decreased likelihood of regression GOLD 2 to GOLD 3-4 but not progression. These findings align with the literature on how smoking impacts lung function decline and symptom exacerbation,^42–44^ suggesting that cessation interventions should start ideally at early stages of the disease as their impact in disease progression decreases as stage severity increases.^45^

Our data suggest that intervening at early stages of airflow limitation may be the most effective strategy, as it is during those stages that individuals have an equal opportunity to regress to a less severe disease stage or progress. Therefore, any preventive measure can be the most effective if implemented alongside early diagnosis.^46^ Unfortunately, COPD is severely underdiagnosed^47^ and when it is diagnosed, diagnosis often comes at very late stages, reducing the potential effect of lifestyle interventions. Early detection represents a challenge, particularly as mean time spent at early stages is less than 7 years and there is a lack of tools for COPD early detection.

### Strengths and Limitations

We did not consider the impact of the use of other nicotine/tobacco products due to the small sample size of individuals reporting e-cigarette use in COPDGene. As other nicotine products use becomes more popular, possibly as cigarette smoking cessation tools,^48^ it will be important to revisit our work to reflect the risk related to these new product use. We were also limited in analyzing NHB and NHW individuals; assessment of other race/ethnicity groups could provide critical information on US-specific health outcomes and health inequalities.

Our study is one of the few to report on transitions among airflow limitation categories across the spectrum of COPD severity stages. Stratifying COPD by stages of severity (GOLD 0, PRISm, GOLD 1, GOLD 2, GOLD 3-4) allowed us to develop detailed insight into the progress of the disease, estimate time spent in various stages of the disease and to characterize the variability of progression within the disease stages. Further, the 10 years follow-up of COPDGene data permits estimating longer-time frame transition probabilities using the Markov Transition modeling framework. Moreover, including relevant risk factors and their analysis enables us to determine better how each factor influences the progression or regression between COPD stages.

## CONCLUSION

Our analysis demonstrates that GOLD 1 and PRISm are the most dynamic COPD stages with the least amount of time being spent in these disease stages than any other. We also found that patient characteristics such as cigarette use history, BMI and previous COPD diagnosis are associated with both disease progression and regression at early stages. This study adds to the evidence that preventive measures such as cessation interventions and prompt detection are key to reducing the COPD burden.

## Data Availability

The COPDGene datasets presented in this study are not readily available. Requests to access these datasets should be directed to COPDGene: https://copdgene.org/research/phase-2-study-documents/

## Acknowledgments

COPDGene Clinical Epidemiology group: for their comments

