## Supplemental Material for "Determinants of COPD Stage Progression and Regression: a Markov Transition Analysis of The COPDGene Cohort"

**SUPPLEMENTARY APPENDIX 1 Sensitivity Analysis**

When defining airflow limitation categories using pre-BD rather than post-BD spirometry values, we observed bigger changes in transition probabilities at GOLD 0 and PRISm. The percentage of individuals that remain at GOLD 0 after five years decreased from 76.2% using post-BD to 72.2 % using pre-BD. Consequently, this increased the percentage of individuals moving from GOLD 0 to PRISm from 7.7% with post-BD to 9.3% using pre-BD and from GOLD 0 to GOLD 2 from 2.8% with post-BD to 3.6% with pre-BD.

The percentage of individuals that remained in the PRISm stage was reduced from 50.3% with post-BD to 47.7% pre-BD). We observed an increase in individuals transitioning from PRISm to GOLD 2 (from 20.4% with post-BD to 25.5% with pre-BD values). The percentage of individuals remaining at GOLD 1 was slightly reduced, with a consequential increase in those who regressed to GOLD 0 (from 17.5% with post-BD to 21.0% with pre-BD) (Figure E1). Further analysis did not show any substantial differences between using pre-BD and post-BD values in the likelihood of transitioning in and out of COPD severity stages (Table E1 and E2).

**Figure E1. One-year (A) and five-year (B) transition probability between PRISm, GOLD levels and Death estimated from all-wave sample with COPD diagnosis using Pre-bronchodilator values.**

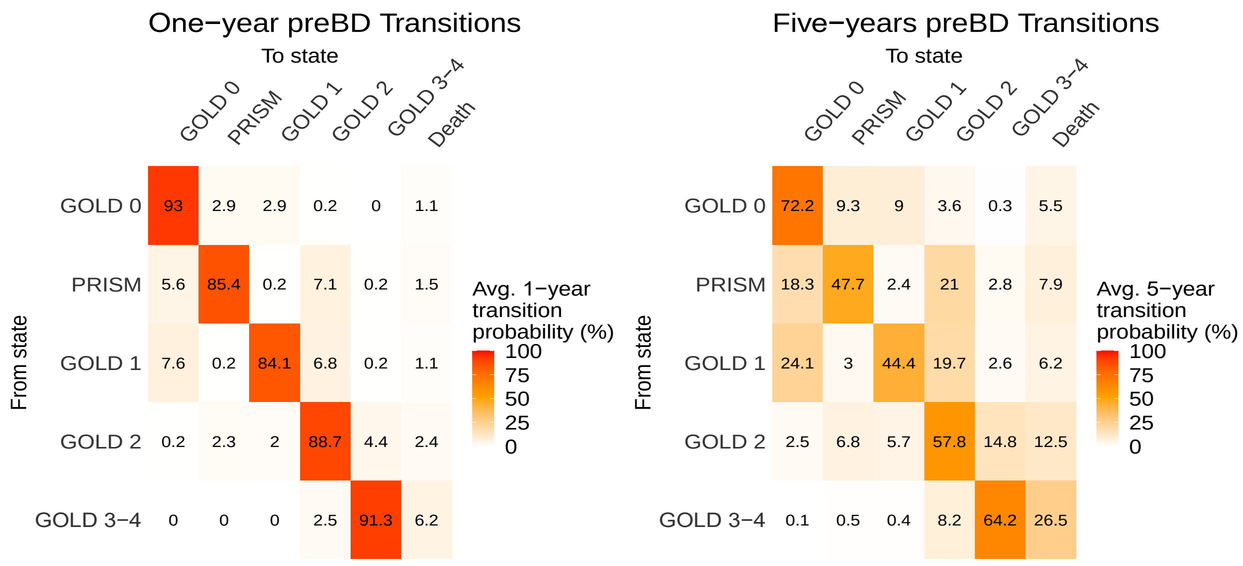

**Table E1. Point estimate and 95% confidence intervals for the transition hazard rates and one-wave transition probabilities of GOLD definition. Results from univariate model using pre-bronchodilator values.**

| | Hazard Rate | | | Probability one year | | |
|---|---|---|---|---|---|---|
| **Transition** | **Estimate** | **95% CI** | | **Estimate** | **95% CI** | |
| **GOLD 0 to** | | **Lower** | **Upper** | | **Lower** | **Upper** |
| GOLD 0 | – | – | – | 92.96 | 92.50 | 93.37 |
| PRISm | 0.032 | 0.029 | 0.036 | 2.86 | 2.55 | 3.20 |
| GOLD 1 | 0.032 | 0.029 | 0.036 | 2.86 | 2.54 | 3.21 |
| GOLD 2 | – | – | – | 0.23 | 0.21 | 0.26 |
| GOLD 3-4 | – | – | – | 0.00 | 0 | 0 |
| Death | 0.011 | 0.009 | 0.013 | 1.08 | 0.93 | 1.27 |
| **PRISm to** | | | | | | |
| GOLD 0 | 0.063 | 0.055 | 0.072 | 5.60 | 4.90 | 6.39 |
| PRISm | – | – | – | 85.42 | 84.33 | 86.46 |
| GOLD 1 | – | – | – | 0.17 | 0.15 | 0.19 |
| GOLD 2 | 0.082 | 0.073 | 0.092 | 7.11 | 6.36 | 7.91 |
| GOLD 3-4 | – | – | – | 0.18 | 0.15 | 0.21 |
| Death | 0.015 | 0.012 | 0.020 | 1.52 | 1.21 | 1.95 |
| **GOLD 1 to** | | | | | | |
| GOLD 0 | 0.086 | 0.075 | 0.100 | 7.63 | 6.68 | 8.79 |
| PRISm | – | – | – | 0.21 | 0.18 | 0.24 |
| GOLD 1 | – | – | – | 84.09 | 82.65 | 85.39 |
| GOLD 2 | 0.079 | 0.068 | 0.091 | 6.82 | 5.89 | 7.74 |
| GOLD 3-4 | – | – | – | 0.17 | 0.15 | 0.20 |
| Death | 0.010 | 0.007 | 0.016 | 1.09 | 0.77 | 1.56 |
| **GOLD 2 to** | | | | | | |
| GOLD 0 | – | – | – | 0.16 | 0.14 | 0.19 |
| PRISm | 0.026 | 0.022 | 0.031 | 2.28 | 1.93 | 2.72 |
| GOLD 1 | 0.023 | 0.019 | 0.027 | 1.97 | 1.64 | 2.37 |
| GOLD 2 | – | – | – | 88.72 | 87.96 | 89.42 |
| GOLD 3-4 | 0.049 | 0.044 | 0.055 | 4.44 | 3.99 | 4.98 |
| Death | 0.024 | 0.020 | 0.028 | 2.43 | 2.10 | 2.78 |
| **GOLD 3-4 to** | | | | | | |
| GOLD 0 | – | – | – | 0.00 | 0.00 | 0.00 |
| PRISM | – | – | – | 0.03 | 0.03 | 0.04 |
| GOLD 1 | – | – | – | 0.03 | 0.02 | 0.04 |
| GOLD 2 | 0.028 | 0.023 | 0.033 | 2.48 | 2.08 | 2.93 |
| GOLD 3-4 | – | – | – | 91.28 | 90.57 | 91.92 |
| Death | 0.064 | 0.058 | 0.071 | 6.18 | 5.66 | 6.79 |

**Table E2. Point estimates and 95% confidence intervals for the transition hazards to regress vs. progress to a more severe COPD GOLD stage for initial COPD severity stages. Results from univariate models using pre-bronchodilator values**

| Demographic | From GOLD 0 to GOLD 1 vs. PRISm (ref) | From PRISm to GOLD 2 vs. GOLD 0 (ref) | From GOLD 1 to GOLD 2 vs. GOLD 0(ref) | From GOLD 1 to GOLD 2 vs. PRISm to GOLD 2 (ref) | From GOLD 2 to GOLD3-4 vs. PRISm (ref) | From GOLD 2 to GOLD 3-4 vs. GOLD 1 (ref) | From GOLD 2 to GOLD 1 vs. PRISm (ref) |
|---|---|---|---|---|---|---|---|
| **Overall** | 1.01 (0.85,1.20) | **1.30 (1.09,1.56)[b]** | 0.91 (0.75,1.12) | 0.97 (0.79,1.18) | **1.89 (1.53,2.32)[b]** | **2.17 (1.75,2.69)[b]** | 0.87 (0.66,1.15) |
| **Gender** | | | | | | | |
| Male | 0.99 (0.77,1.26) | **1.60 (1.22,2.10)[b]** | 1.07 (0.82,1.41) | 0.84 (0.65,1.10) | **2.09 (1.55,2.82)[b]** | **2.33 (1.72,3.15)[b]** | 0.90 (0.61,1.33) |
| Female | 1.03 (0.81,1.31) | 1.11 (0.87,1.42) | 0.74 (0.54,1.00) | 1.06 (0.79,1.43) | **1.71 (1.27,2.29)[b]** | **2.00 (1.47,2.72)[b]** | 0.85 (0.58,1.26) |
| **Age group (years)** | | | | | | | |
| 39 to 54 | 0.88 (0.58,1.35) | 1.24 (0.79,1.94) | 0.56 (0.29,1.09) | 1.74 (0.93,3.25) | 1.40 (0.75,2.61) | **2.36 (1.09,5.08)[b]** | 0.59 (0.23,1.54) |
| 55 to 64 | 1.03 (0.82,1.28) | 1.10 (0.87,1.40) | 0.85 (0.65,1.11) | 1.13 (0.87,1.46) | **1.88 (1.43,2.47)[b]** | **2.45 (1.80,3.34)[b]** | 0.77 (0.52,1.13) |
| 65 to 90 | **1.51 (1.01,2.27)[b]** | **2.04 (1.39,3.00)[b]** | 1.16 (0.81,1.67) | **0.45 (0.32,0.64)[a]** | **1.79 (1.22,2.62)[b]** | **1.78 (1.27,2.50)[b]** | 1.00 (0.64,1.57) |
| **Race** | | | | | | | |
| Non-Hispanic White | 1.17 (0.94,1.46) | **1.57 (1.25,1.97)[b]** | 0.89 (0.71,1.13) | **0.76 (0.61,0.96)[a]** | **1.69 (1.32,2.16)[b]** | **1.88 (1.47,2.40)[b]** | 0.90 (0.66,1.23) |
| Non-Hispanic Black | 0.84 (0.64,1.12) | 0.92 (0.67,1.25) | 1.00 (0.64,1.56) | **1.60 (1.09,2.36)[b]** | **2.33 (1.56,3.47)[b]** | **3.23 (2.03,5.14)[b]** | 0.72 (0.40,1.31) |
| **Cigarrette use** | | | | | | | |
| Current | 0.94 (0.75,1.19) | 1.22 (0.95,1.56) | **0.71 (0.52,0.98)[a]** | 1.28 (0.98,1.67) | **1.86 (1.39,2.50)[b]** | **3.21 (2.21,4.68)[b]** | **0.58 (0.37,0.91)[a]** |
| Former | 1.10 (0.85,1.42) | **1.34 (1.04,1.75)[b]** | 1.52 (1.15,2.02) | **0.72 (0.54,0.97)[a]** | **1.85 (1.38,2.47)[b]** | **1.70 (1.30,2.22)[b]** | 1.09 (0.76,1.55) |

| | | | | | | | |
|---|---|---|---|---|---|---|---|
| **Cigarrette pack years** | | | | | | | |
| less than 20 | 1.17 (0.77,1.78) | 0.93 (0.60,1.46) | **0.35 (0.18,0.70)**[a] | 0.71 (0.36,1.39) | 1.90 (0.97,3.74) | 1.83 (0.97,3.45) | 1.04 (0.46,2.36) |
| 20 or more | 0.98 (0.81,1.18) | 1.40 (1.15,1.71) | 1.02 (0.82,1.27) | 0.96 (0.78,1.18) | **1.87 (1.50,2.33)**[b] | **2.22 (1.76,2.79)**[b] | 0.84 (0.63,1.13) |
| **Body Mass Index (kg/m2)** | | | | | | | |
| Normal (≤ 24.9 ) | **2.23 (1.56,3.19)**[b] | **1.81 (1.11,2.93)**[b] | 0.99 (0.71,1.37) | 1.24 (0.85,1.82) | **2.83 (1.78,4.50)**[b] | **2.79 (1.80,4.31 )**[b] | 1.01 (0.55,1.88) |
| Overweight (25 to 29.9) | 1.27 (0.95,1.69) | 1.32 (0.94,1.85) | 1.02 (0.73,1.43) | 1.20 (0.86,1.67) | **1.83 (1.27,2.63)**[b] | **1.78 (1.24,2.54)**[b] | 1.03 (0.65,1.62) |
| Obesity (≥30) | **0.47 (0.35,0.65)**[a] | 1.21 (0.95,1.54) | **0.64 (0.42,0.98)**[a] | 0.93 (0.63,1.38) | **1.52 (1.11,2.08)**[b] | **2.08 (1.48,2.93)**[b] | 0.73 (0.48,1.12) |
| **COPD diagnosis** | | | | | | | |
| No | 1.08 (0.90,1.29) | 1.05 (0.85,1.28) | 0.81 (0.65,1.02) | 1.07 (0.85,1.34) | 1.04 (0.80,1.36) | **1.35 (1.01,1.80)**[b] | 0.77 (0.55,1.08) |
| Yes | 0.58 (0.30,1.11) | 1.05 (0.85,1.28) | **1.74 (1.01,3.00)**[b] | 0.71 (0.47,1.07) | **4.17 (2.81,6.19)**[b] | NA | 1.06 (0.63,1.80) |

a: Statistical significant hazard ratio <1 (Regression to a less severe stage is more likely)

b: Statistical significant hazard ratio ≥1 (Progression to a more severe stage is more likely)

### SUPPLEMENTARY APPENDIX 2

**Figure E2**. **Schematic depiction of COPDGene participants' trajectories between PRISm and COPD-GOLD severity stages across three Phases***

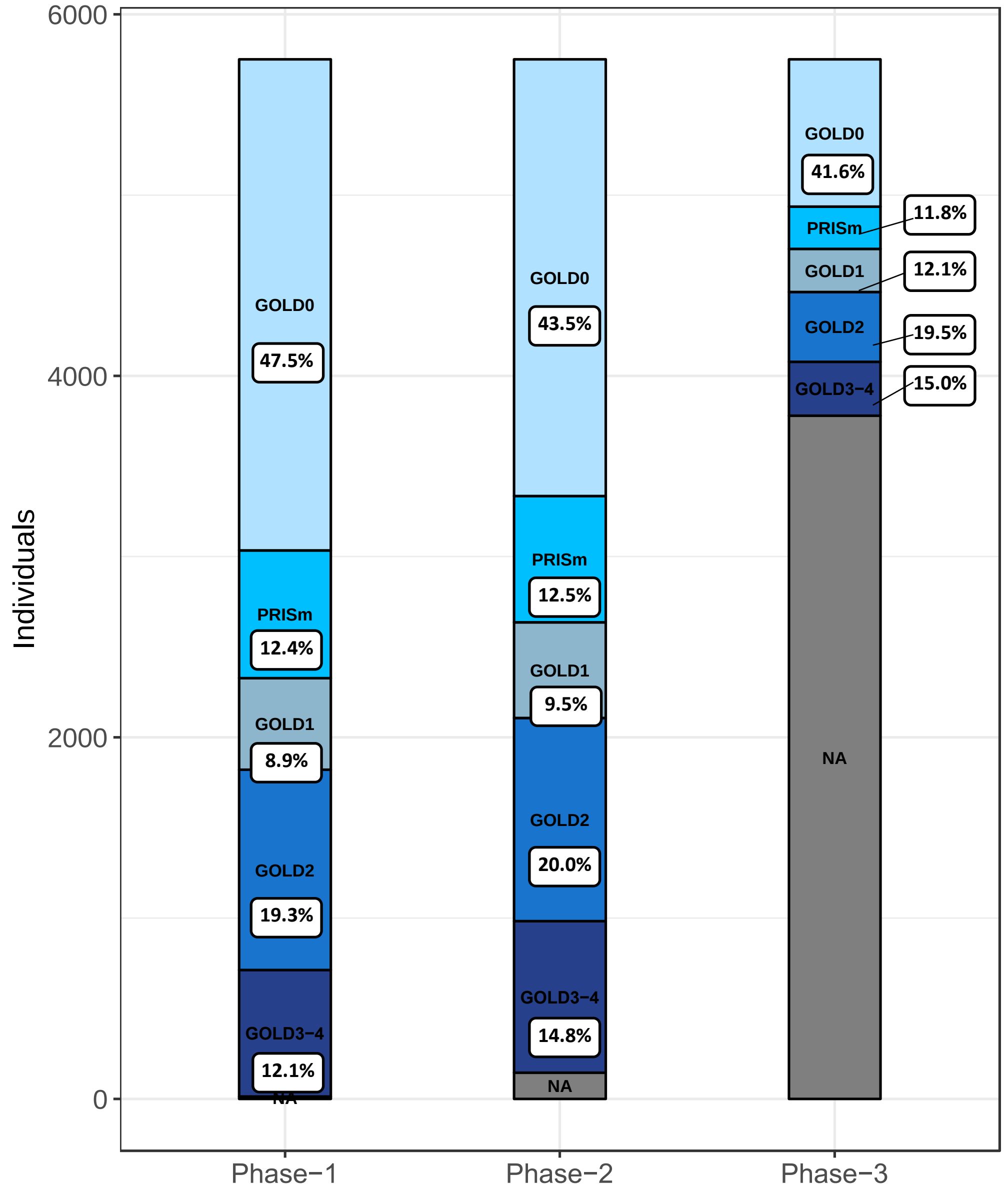


*Each panel represents the number of individuals (proportion %) at each PRISm and GOLD severity stage for each of the COPDGene cohort Phases.
NA: Represents those individuals who were lost at follow-up.

**Table E3. Point estimate and 95% confidence intervals for the transition hazard rates and one-wave transition probabilities. Univariate model**

| | Hazard Rate | | | Probability one year | | |
|---|---|---|---|---|---|---|
| Transition | **Estimate** | **95% CI** | | **Estimate** | **95% CI** | |
| **GOLD 0 to** | | Lower | Upper | | Lower | Upper |
| GOLD 0 | – | – | – | 94.17 | 93.75 | 94.54 |
| PRISm | 0.025 | 0.022 | 0.028 | 2.24 | 2.00 | 2.51 |
| GOLD 1 | 0.027 | 0.025 | 0.031 | 2.45 | 2.20 | 2.73 |
| GOLD 2 | – | – | – | 0.17 | 0.15 | 0.19 |
| GOLD 3-4 | – | – | – | 0.00 | 0.00 | 0.00 |
| Death | 0.009 | 0.008 | 0.011 | 0.96 | 0.83 | 1.11 |
| **PRISm to** | | | | | | |
| GOLD 0 | 0.061 | 0.053 | 0.070 | 5.49 | 4.74 | 6.24 |
| PRISm | – | – | – | 86.52 | 85.32 | 87.57 |
| GOLD 1 | – | – | – | 0.13 | 0.12 | 0.15 |
| GOLD 2 | 0.066 | 0.058 | 0.076 | 5.80 | 5.13 | 6.61 |
| GOLD 3-4 | – | – | – | 0.15 | 0.12 | 0.17 |
| Death | 0.019 | 0.015 | 0.024 | 1.91 | 1.54 | 2.42 |
| **GOLD 1 to** | | | | | | |
| GOLD 0 | 0.070 | 0.060 | 0.082 | 6.29 | 5.43 | 7.24 |
| PRISm | – | – | – | 0.17 | 0.15 | 0.19 |
| GOLD 1 | – | – | – | 84.81 | 83.35 | 86.05 |
| GOLD 2 | 0.081 | 0.071 | 0.093 | 7.02 | 6.18 | 7.99 |
| GOLD 3-4 | – | – | – | 0.18 | 0.15 | 0.21 |
| Death | 0.015 | 0.011 | 0.021 | 1.53 | 1.16 | 2.05 |
| **GOLD 2 to** | | | | | | |
| GOLD 0 | – | – | – | 0.14 | 0.12 | 0.16 |
| PRISm | 0.026 | 0.022 | 0.031 | 2.30 | 1.92 | 2.72 |
| GOLD 1 | 0.021 | 0.017 | 0.026 | 1.80 | 1.48 | 2.22 |
| GOLD 2 | – | – | – | 88.56 | 87.75 | 89.30 |
| GOLD 3-4 | 0.050 | 0.045 | 0.056 | 4.48 | 4.01 | 5.01 |
| Death | 0.027 | 0.023 | 0.031 | 2.71 | 2.34 | 3.10 |
| **GOLD 3-4 to** | | | | | | |
| GOLD 0 | – | – | – | 0.00 | 0.00 | 0.00 |
| PRISM | – | – | – | 0.03 | 0.03 | 0.04 |
| GOLD 1 | – | – | – | 0.03 | 0.02 | 0.03 |
| GOLD 2 | 0.029 | 0.024 | 0.034 | 2.56 | 2.13 | 3.06 |
| GOLD 3-4 | – | – | – | 91.00 | 90.18 | 91.69 |
| Death | 0.066 | 0.060 | 0.074 | 6.38 | 5.80 | 7.03 |

**Figures E3 (1-7). Five-years transition probabilities between COPD-GOLD severity stages by sociodemographic characteristics**

**Figure E3-1.-Five-year transition probability between COPD severity levels estimated for a) Males and b) Females**

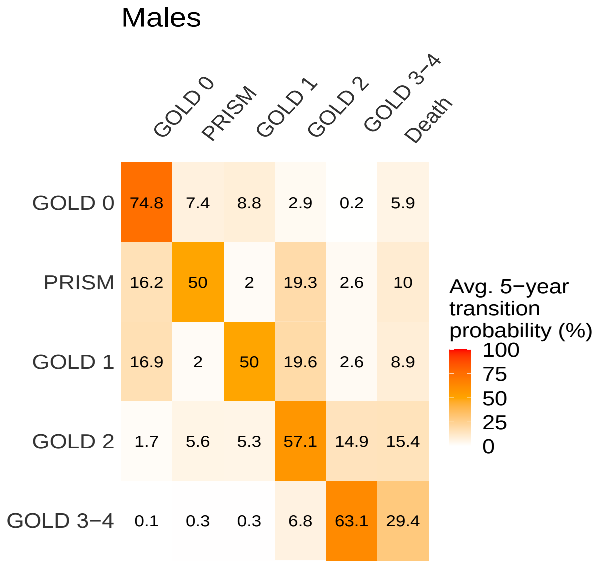


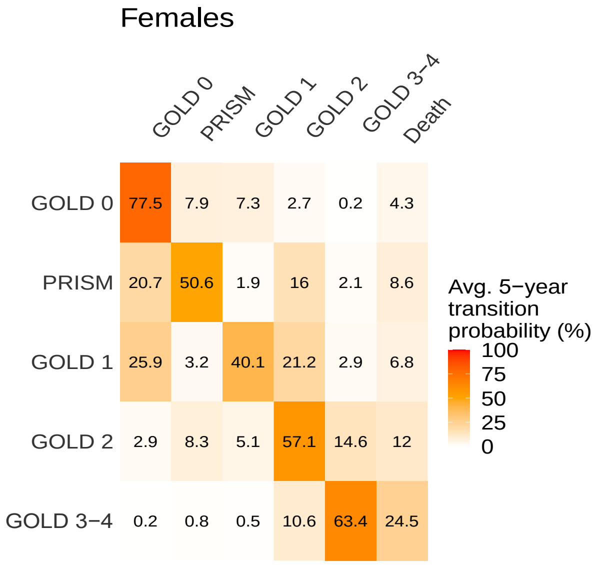


**Figure E3-2. A Five-year transition probability between COPD-GOLD severity levels estimated for a) 30 to 49 years old b) 50 to 64 years old and c) 65 to 90 years old**

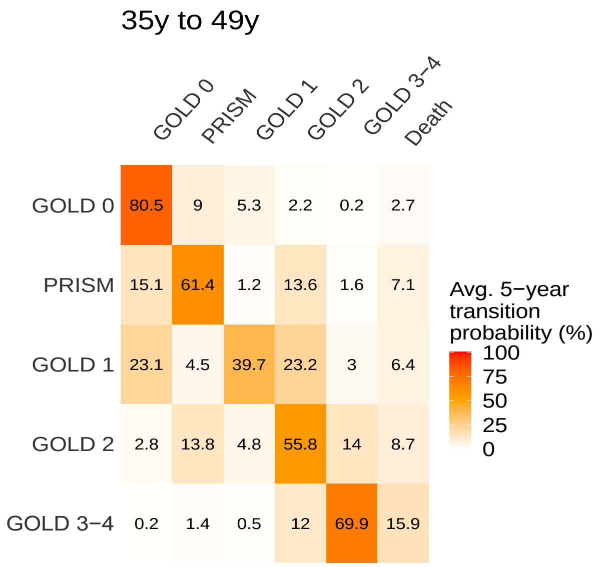


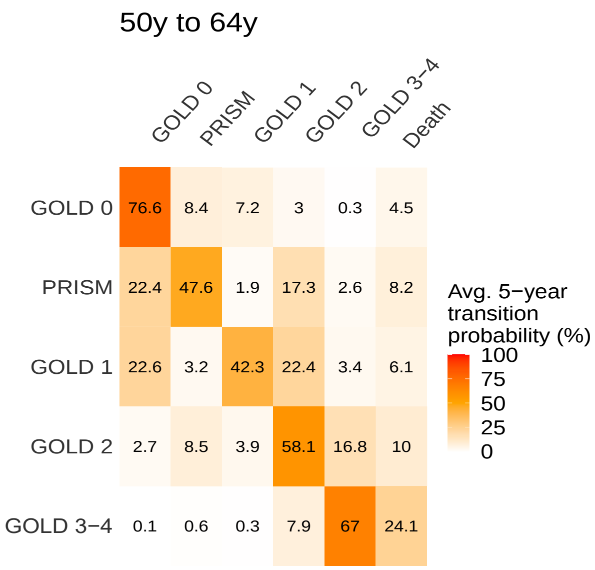

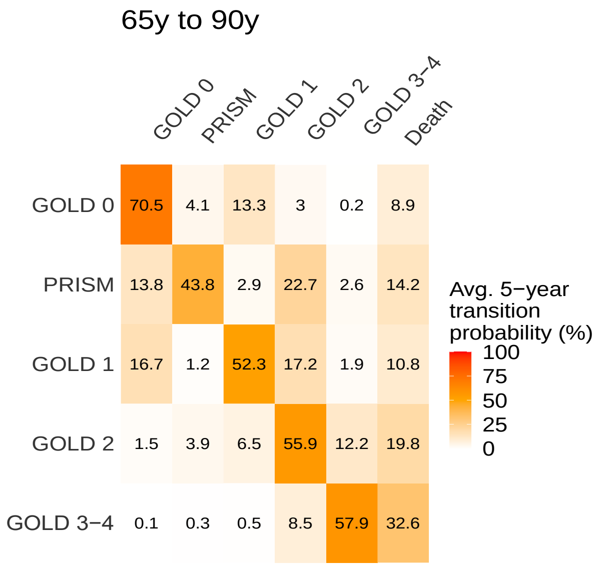


**Figure E3-3. Five-year transition probability between COPD severity levels estimated for a) Non-Hispanic White and b) Non-Hispanic Black.**

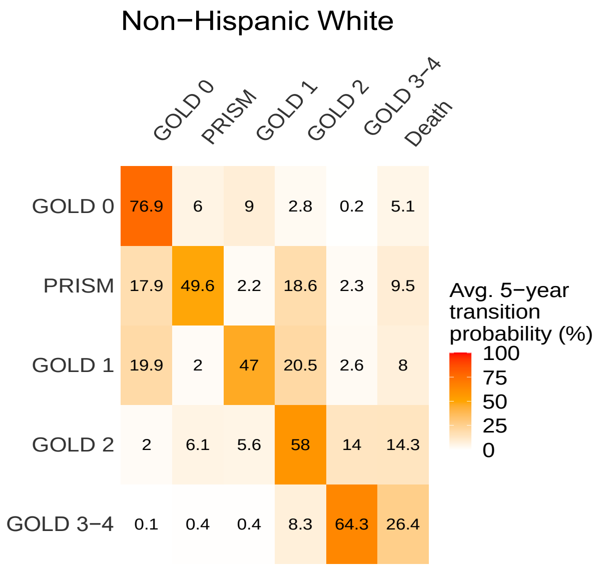


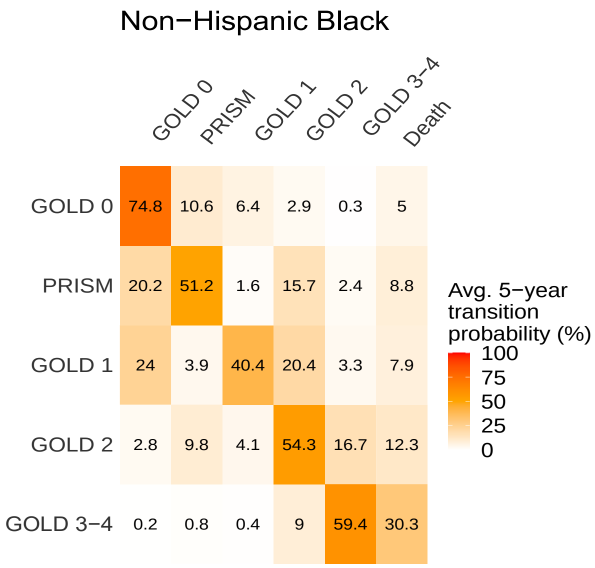

**Figure E3-4. Five-year transition probability between COPD severity levels estimated for a) Current smokers and b) Former smokers.**

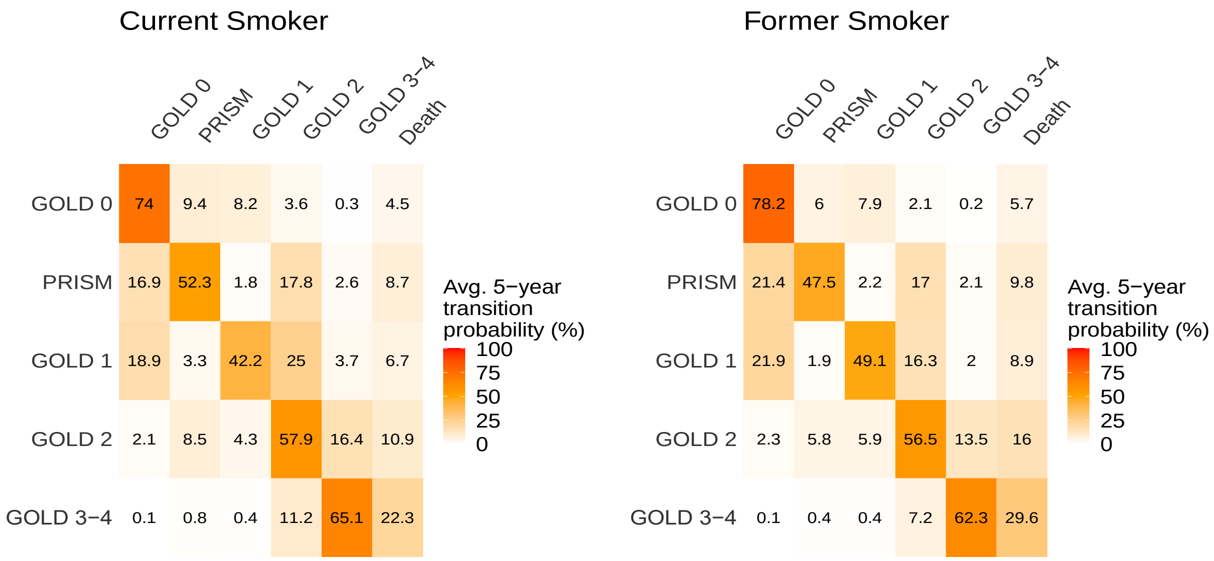


**Figure E3-5. Five-year transition probability between COPD severity levels estimated for number of pack of cigarettes per years a) less than 20 and b) 20 or more.**

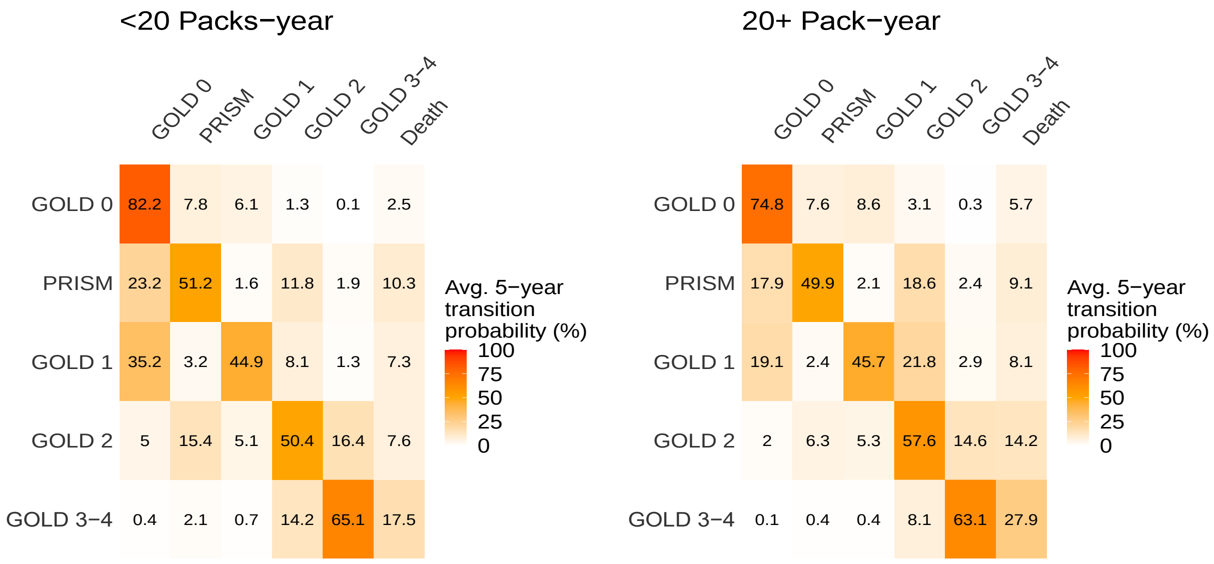

**Figure E3-6. Five-year transition probability between COPD severity levels estimated for three categories of BMI a) Normal b) Overweight and c) Obesity**

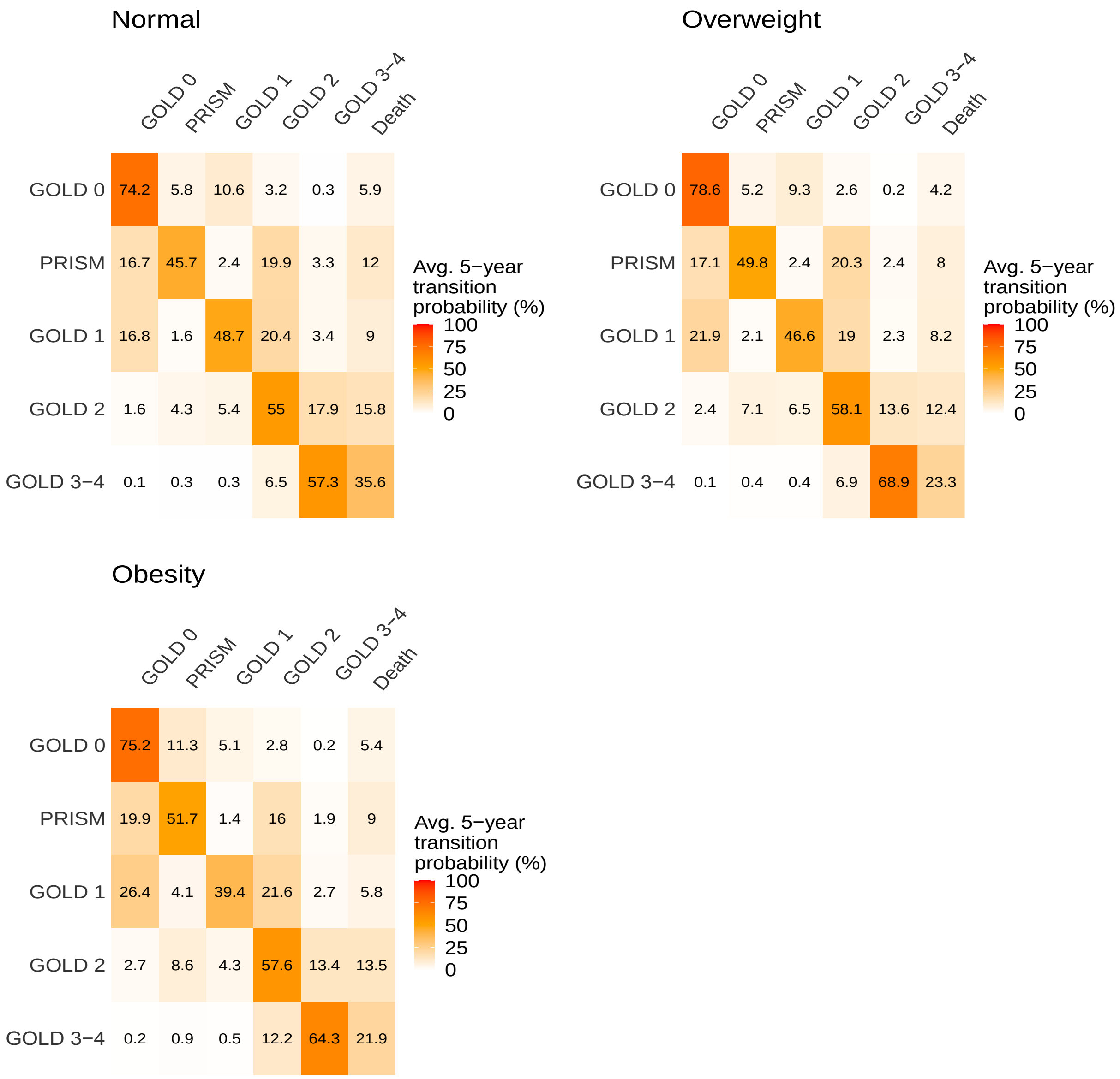

**Figure E3-7. Five-year transition probability between COPD severity levels estimated for a) No COPD previous diagnosis and b) COPD previous diagnosis.**

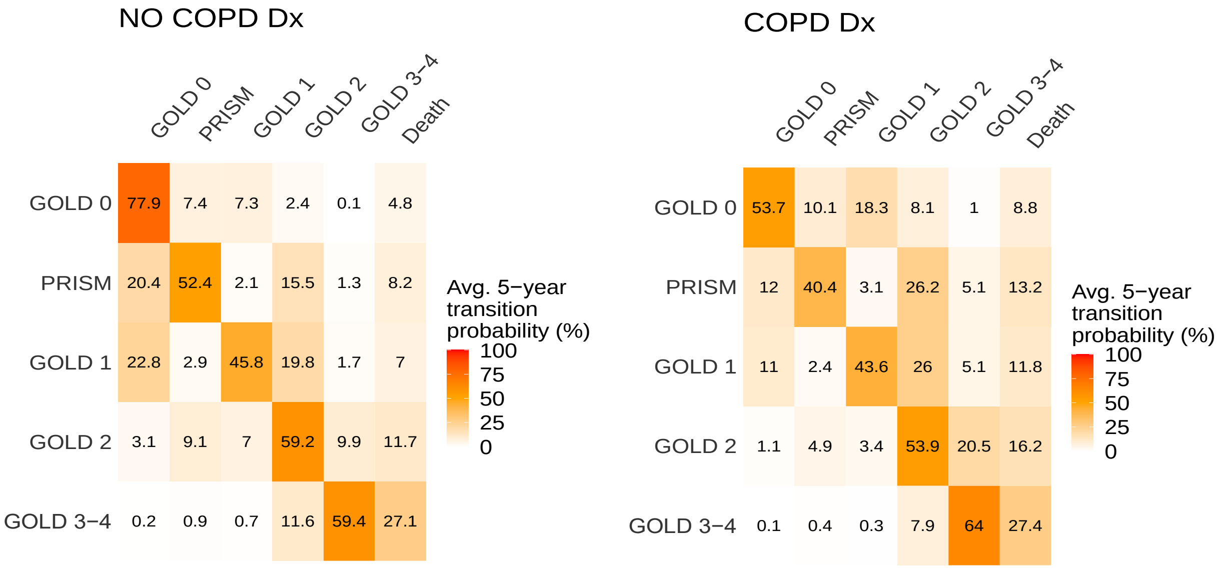

**Table E4. Point estimates and 95% confidence intervals for the transition hazards to progress to a more severe GOLD stage by sociodemographic characteristics. Results from the univariate model**

| Demographic | GOLD 0 to PRISm | GOLD 0 to GOLD 1 | PRISm to GOLD 2 | GOLD 1 to GOLD 2 | GOLD 2 to GOLD 3-4 |
|---|---|---|---|---|---|
| **Gender** | | | | | |
| Male | 1 (Ref.) | 1 (Ref.) | 1 (Ref.) | 1 (Ref.) | 1 (Ref.) |
| Female | 1.05(0.83,1.32) | 0.90(0.72,1.13) | 0.82(0.63,1.08) | 1.21(0.92,1.59) | 0.98(0.78,1.22) |
| **Age group (years)** | | | | | |
| 39 to 54 | 0.92(0.70,1.21) | 0.73(0.52,1.02) | 0.71(0.49,1.03) | 1.10(0.70,1.72) | 0.84(0.56,1.25) |
| 55 to 64 | 1 (Ref.) | 1 (Ref.) | 1 (Ref.) | 1 (Ref.) | 1 (Ref.) |
| 65 to 90 | **0.52(0.36,0.75)**[a] | **1.75(1.38,2.24)**[b] | **1.39(1.00,1.92)**[b] | **0.71(0.53,0.95)**[a] | 0.80(0.62,1.02) |
| **Race** | | | | | |
| NH-White | 1 (Ref.) | 1 (Ref.) | 1 (Ref.) | 1 (Ref.) | 1 (Ref.) |
| NH-Black | **1.79(1.42,2.26)**[b] | **0.77(0.60,0.99)** | 0.87(0.66,1.15) | 1.10(0.79,1.54) | **1.29(1.01,1.66)**[b] |
| **Cigarrette use** | | | | | |
| Current | 1 (Ref.) | 1 (Ref.) | 1 (Ref.) | 1 (Ref.) | 1 (Ref.) |
| Former | **0.64(0.51,0.81)**[a] | 0.86(0.69,1.08) | 1.01(0.77,1.33) | **0.61(0.46,0.80)**[a] | 0.84(0.67,1.06) |
| **Cigarrette pack years** | | | | | |
| less than 20 | 1 (Ref.) | 1 (Ref.) | 1 (Ref.) | 1 (Ref.) | 1 (Ref.) |
| 20 or more | 1.04(0.77,1.39) | **1.47(1.05,2.06)**[b] | 1.46(0.95,2.26) | **2.52(1.24,5.11)**[b] | 0.84(0.54,1.29) |
| **Body Mass Index (kg/m2)** | | | | | |
| Normal (≤24.9 ) | 1 (Ref.) | 1 (Ref.) | 1 (Ref.) | 1 (Ref.) | 1 (Ref.) |
| Overweight (25 to 29.9) | 0.82(0.58,1.16) | 0.87(0.67,1.12) | 0.96(0.65,1.41) | 0.93(0.68,1.26) | **0.67(0.51,0.88)**[a] |
| Obesity (≥30) | **1.85(1.37,2.49)**[b] | 0.53(0.39,0.71) | 0.75(0.52,1.07) | 1.14(0.79,1.65) | **0.69(0.53,0.91)**[a] |
| **COPD diagnosis** | | | | | |
| No | 1 (Ref.) | 1 (Ref.) | 1 (Ref.) | 1 (Ref.) | 1 (Ref.) |
| Yes | **1.88(1.25,2.84)**[b] | **3.17(2.34,4.30)**[b] | **2.01(1.49,2.72)**[b] | **1.41(1.03,1.94)**[b] | **2.11(1.67,2.67)**[b] |

NH-White: Non-Hispanic White; NH-Black- Non-Hispanic Black

[a]: Statistical significant hazard ratio <1 (Less likely to progress to a more severe stage)

[b]: Statistical significant hazard ratio ≥1 (More likely to progress to a more severe stage)

**Table E5. Point estimates and 95% confidence intervals for the transition hazards to regress to a less severe GOLD stage by sociodemographic characteristics.**

| Demographic | PRISm to GOLD 0 | GOLD 1 to GOLD 0 | GOLD 2 to PRISm | GOLD 2 to GOLD 1 | GOLD 3-4 to GOLD 2 |
|---|---|---|---|---|---|
| **Gender** | | | | | |
| Male | 1 (Ref.) | 1 (Ref.) | 1 (Ref.) | 1 (Ref.) | 1 (Ref.) |
| Female | 1.26(0.94,1.68) | **1.67(1.22,2.29)**[b] | **1.46(1.02,2.09)**[b] | 1.07(0.72,1.61) | **1.58(1.09,2.27)**[b] |
| **Age group (years)** | | | | | |
| 39 to 54 | **0.57(0.39,0.84)**[a] | 1.02(0.57,1.82) | 1.46(0.89,2.38) | 1.34(0.59,3.01) | 1.52(0.76,3.01) |
| 55 to 64 | 1 (Ref.) | 1 (Ref.) | 1 (Ref.) | 1 (Ref.) | 1 (Ref.) |
| 65 to 90 | **0.66(0.44,0.97)**[a] | **0.70(0.50,0.99)**[a] | **0.49(0.32,0.74)**[a] | **1.58(1.01,2.47)**[b] | 1.18(0.81,1.72) |
| **Race** | | | | | |
| NH-White | 1 (Ref.) | 1 (Ref.) | 1 (Ref.) | 1 (Ref.) | 1 (Ref.) |
| NH-Black | 1.14(0.86,1.53) | 1.31(0.89,1.93) | **1.65(1.14,2.40)**[b] | 0.81(0.48,1.36) | 1.17(0.75,1.81) |
| **Cigarrette use** | | | | | |
| Current | 1 (Ref.) | 1 (Ref.) | 1 (Ref.) | 1 (Ref.) | 1 (Ref.) |
| Former | 1.28(0.97,1.70) | 1.05(0.76,1.45) | 0.72(0.51,1.03) | 1.28(0.83,1.98) | **0.66(0.46,0.96)**[a] |
| **Cigarrette pack years** | | | | | |
| less than 20 | 1 (Ref.) | 1 (Ref.) | 1 (Ref.) | 1 (Ref.) | 1 (Ref.) |
| 20 or more | 0.82(0.57,1.18) | 0.57(0.37,0.88) [a] | 0.38(0.23,0.61) [a] | 0.97(0.44,2.16) | 0.53(0.29,0.97)[a] |
| **Body Mass Index (kg/m2)** | | | | | |
| Normal (≤24.9 ) | 1 (Ref.) | 1 (Ref.) | 1 (Ref.) | 1 (Ref.) | 1 (Ref.) |
| Overweight (25 to 29.9) | 0.95(0.59,1.53) | 1.29(0.88,1.88) | 1.54(0.91,2.60) | 1.19(0.72,1.96) | 0.95(0.58,1.55) |
| Obesity (≥30) | 1.13(0.75,1.71) | **1.72(1.14,2.57)**[b] | **1.82(1.12,2.97)**[b] | 0.86(0.51,1.43) | **1.75(1.13,2.72)**[b] |
| **COPD diagnosis** | | | | | |
| No | 1 (Ref.) | 1 (Ref.) | 1 (Ref.) | 1 (Ref.) | 1 (Ref.) |
| Yes | 0.82(0.54,1.23) | 0.61(0.35,1.06) | **0.65(0.45,0.95)**[a] | **0.51(0.32,0.81)**[a] | 0.68(0.45,1.04) |

NH-White: Non-Hispanic White; NH-Black- Non-Hispanic Black

[a]: Statistical significant hazard ratio <1 (Less likely to regress to a less severe stage)

[b]: Statistical significant hazard ratio ≥1 (More likely to regress to a less severe stage)

**Figure E4. HRs by sociodemographic characteristics for transitions corresponding to COPD progression to Death.**

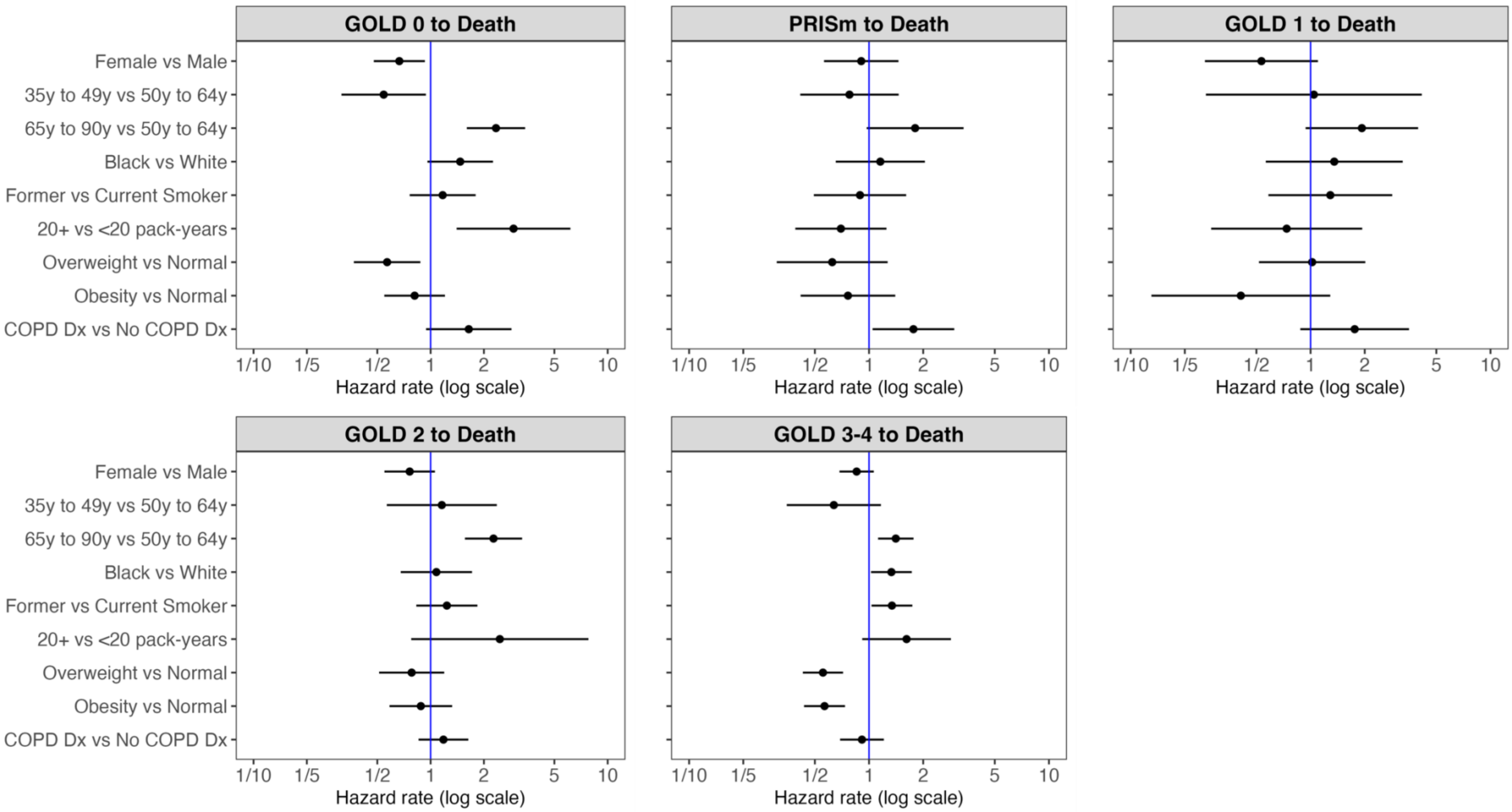

**Table E6. Point estimates and 95% confidence intervals for the transition hazards to progress to Death by sociodemographic characteristics.**

| Demographic | PRISm to GOLD 0 | GOLD 1 to GOLD 0 | GOLD 2 to PRISm | GOLD 2 to GOLD 1 | GOLD 3-4 to GOLD 2 |
|---|---|---|---|---|---|
| **Gender** | | | | | |
| Male | 1 (Ref.) | 1 (Ref.) | 1 (Ref.) | 1 (Ref.) | 1 (Ref.) |
| Female | **0.70(0.50,0.97)**[a] | 0.94(0.58,1.51) | 0.76(0.39,1.48) | 0.76(0.55,1.05) | 0.82(0.66,1.02) |
| **Age group (years)** | | | | | |
| 39 to 54 | **0.54(0.31,0.93)** [a] | 0.85(0.47,1.55) | 1.22(0.38,3.96) | 1.00(0.48,2.07) | 0.63(0.35,1.15) |
| 55 to 64 | 1 (Ref.) | 1 (Ref.) | 1 (Ref.) | 1 (Ref.) | 1 (Ref.) |
| 65 to 90 | **2.09(1.48,2.96)** [b] | 1.60(0.90,2.84) | 1.75(0.89,3.46) | **2.51(1.77,3.57)** [b] | **1.42(1.15,1.76)** [b] |
| **Race** | | | | | |
| White | 1 (Ref.) | 1 (Ref.) | 1 (Ref.) | 1 (Ref.) | 1 (Ref.) |
| Black | 0.96(0.68,1.35) | 0.96(0.59,1.56) | 1.08(0.50,2.33) | 0.71(0.46,1.09) | 1.20(0.94,1.53) |
| **Cigarrette use** | | | | | |
| Current | 1 (Ref.) | 1 (Ref.) | 1 (Ref.) | 1 (Ref.) | 1 (Ref.) |
| Former | 1.34(0.96,1.88) | 1.06(0.65,1.71) | 1.39(0.72,2.67) | **1.65(1.17,2.31)** [b] | **1.36(1.07,1.73)** [b] |
| **Cigarrette pack years** | | | | | |
| less than 20 | 1 (Ref.) | 1 (Ref.) | 1 (Ref.) | 1 (Ref.) | 1 (Ref.) |
| 20 or more | **3.31(1.55,7.07)**[b] | 0.69(0.39,1.23) | 0.79(0.31,2.03) | 2.54(0.73,8.83) | 1.63(0.92,2.89) |
| **Body Mass Index (kg/m2)** | | | | | |
| Normal (≤24.9 ) | 1 (Ref.) | 1 (Ref.) | 1 (Ref.) | 1 (Ref.) | 1 (Ref.) |
| Overweight (25 to 29.9) | 0.70(0.45,1.08) | 0.60(0.29,1.24) | 1.00(0.51,1.94) | 0.92(0.60,1.41) | **0.60(0.46,0.76)** [a] |
| Obesity (≥30) | 0.95(0.64,1.41) | 0.71(0.39,1.29) | 0.47(0.15,1.47) | 1.05(0.71,1.57) | **0.56(0.43,0.73)** [a] |
| **COPD diagnosis** | | | | | |
| No | 1 (Ref.) | 1 (Ref.) | 1 (Ref.) | 1 (Ref.) | 1 (Ref.) |
| Yes | 1.75(0.99,3.10) | 1.60(0.93,2.77) | 1.76(0.89,3.49) | 1.33(0.96,1.84) | 0.98(0.75,1.29) |

[a]: Statistical significant hazard ratio <1 (Less likely to progress to death)

[b]: Statistical significant hazard ratio ≥1 (More likely to progress to death)

**Figure E4**. **Schematic depiction of COPDGene participants with three observations' trajectories between PRISm and COPD-GOLD severity stages across three Phases.**

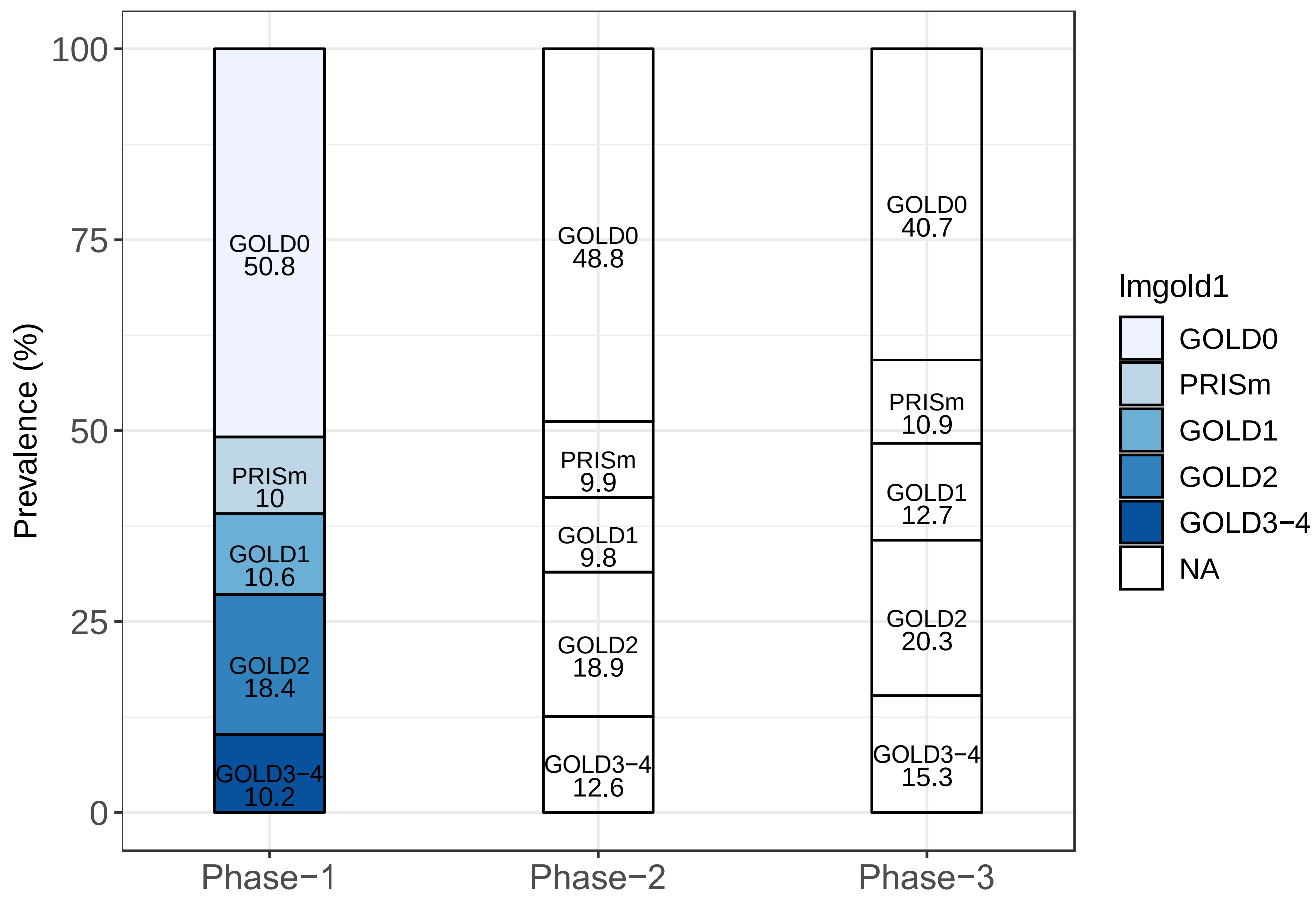

**Figure E5. One-year (A) and five-year (B) transition probability between PRISm, GOLD levels and Death estimated from all-wave sample with COPD diagnosis using complete-case analysis.**

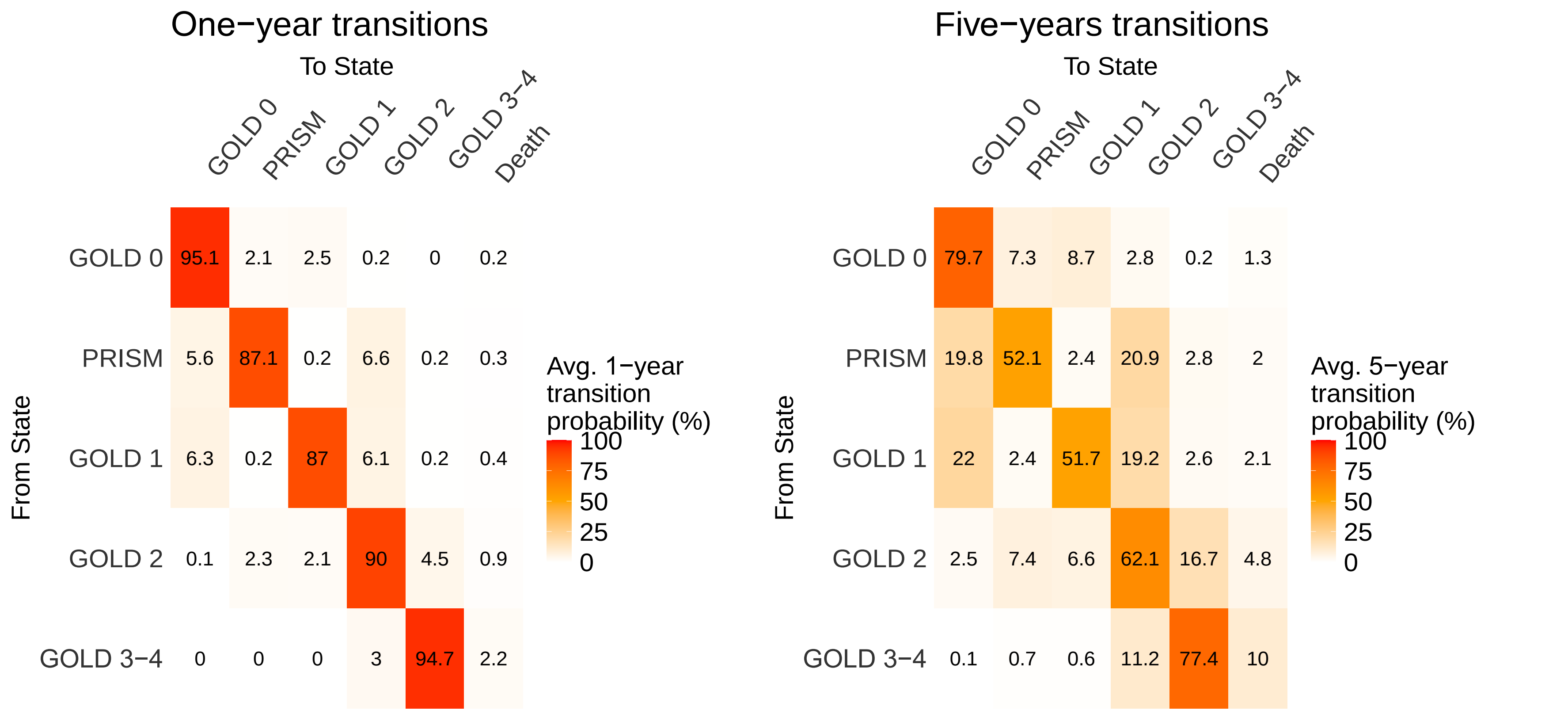
